# Quantifying variant contributions in cystic kidney disease using national-scale whole genome sequencing

**DOI:** 10.1101/2024.02.14.24302377

**Authors:** Omid Sadeghi-Alavijeh, Melanie MY Chan, Gabriel Doctor, Catalin D Voinescu, Alexander Stuckey, Athanasios Kousathanas, Alexander Ho, Genomics England Research Consortium, Horia C Stanescu, Detlef Bockenhauer, Richard N Sandford, Adam P Levine, Daniel P Gale

**Author notes:** Corresponding author: Daniel P Gale.

## Abstract

Cystic kidney disease (CyKD) is a predominantly familial disease. Gene discovery has been led by family-based and candidate gene studies, limiting the ascertainment of additional genome-wide variants involved. Taking whole genome sequencing data from the 100,000 Genomes Project, we used hypothesis-free approaches to systematically characterize and quantify genetic contributors to CyKD across variant types and the allelic spectrum in 1,209 cases and 26,096 ancestry-matched controls. In 82.3% CyKD cases a likely disease-causing monogenic variant was identified. There was an enrichment of rare coding, splicing and structural variants in known CyKD genes, and novel gene-based signals in *COL4A3* and (monoallelic) *PKHD1*. The risks of these variants to CyKD were quantified, with lower risk seen in more recently described genes. Meta-analysis of common variants across four international biobanks did not reveal any associations. Common variants accounted for 3-9% of the heritability of CyKD across three European ancestry cohorts. This represents an unbiased examination of the genetic architecture of a national CyKD cohort using robust statistical methodology. We have quantified the contribution of coding, non-coding, and structural variants to CyKD, and the small contribution of common variants to its heritability. These findings will inform genetic testing and counselling strategies in the clinic.

## Introduction

Cystic kidney disease (CyKD) is a term for any disease that causes multiple fluid filled cysts within the kidney (excluding cystic degeneration of chronically diseased/failed kidneys). The term Polycystic Kidney Disease (PKD) generally refers to CyKD in which the kidneys become enlarged and replaced by cysts and is almost always inherited as a Mendelian trait (either autosomal dominant or autosomal recessive). 90% of CyKD is caused by Autosomal Dominant Polycystic Kidney Disease (ADPKD) which accounts for approximately 10% of all patients receiving kidney replacement therapy for kidney failure (KF) in the United Kingdom (UK)^1^. It is estimated that between 1 in 800-1,000 of the population has ADPKD^2^ making this the commonest monogenic cause of life-shortening disease worldwide.

Rare monoallelic variants in two genes, *PKD1* and *PKD2,* cause the majority of ADPKD, whilst biallelic variants in *PKHD1* cause the majority of ARPKD. *PKD1* accounts for ∼80% of ADPKD diagnoses whilst *PKD2* accounts for ∼15%^2^. *PKD1* encodes Polycystin-1 (PC1), a large multi-domain glycoprotein whilst *PKD2* encodes Polycstin-2 (PC2) which is a nonspecific cation channel that interacts with PC1. Both proteins are found in primary cilia and play a role in mechanotransduction, transferring external information to the cell. Whilst the function of PC1, PC2 and the PC1-2 complex is still not fully understood, it is increasingly accepted that these proteins prevent cystogenesis via a dose dependent mechanism^3^. *PKHD1* encodes the fibrocystin protein and acts via a common polycystin pathway to cause cysts in ARPKD usually manifesting at younger age than ADPKD and associated with extra-renal symptoms including liver fibrosis^4^.

There has been increasing recognition of the genetic heterogeneity of CyKD including contributions of genes other than *PKD1/PKD2*. Among the 10-15% of patients suspected to have ADPKD who have no explaining variants in either *PKD1* or *PKD2,* next generation sequencing (NGS) has identified several other genes in which variants cause ADPKD including *DNAJB11*^5^*, GANAB*^6^*, ALG9*^7^ and more recently *IFT140*^8^ and *ALG5*^9^. Presently, a clinical diagnosis of polycystic kidney disease implies a high risk of kidney failure but pathogenic variants in these more recently described genes seem to carry a lower penetrance, manifesting as sporadic or non-Mendelian disease in individuals or families, with a less severe phenotype. To date, estimates of penetrance and outcomes with these rarer genetic variants are based predominantly on case series and reports of individual families, increasing the risk of ascertainment bias because more severely affected families are more likely to have come to light. Providing accurate data about the adverse consequences of these rarer variants is essential to prognostication, informing life and reproductive choices for patients and family members.

Obtaining a molecular diagnosis for CyKD allows better prognostication and can inform choice of therapy^10,11^. Genetic testing for *PKD1* variants is challenging owing to its high GC content, numerous repetitive regions, and the presence of six pseudogenes that share 97% of their sequence with *PKD1*^12,13^. This has traditionally necessitated the use of Sanger sequencing of a long-range PCR-amplification product^13^, a method that is technically difficult and expensive. Whole genome sequencing (WGS) has been shown to offer high quality *PKD1* sequencing^14^. WGS provides more uniform coverage and avoids capture bias, resulting in increased sensitivity to detect structural variants and rare single nucleotide variants (SNVs) compared with exome (or capture-based) sequencing platforms, even within regions targeted by the latter^15,16^. In addition, WGS benefits from economies of scale and the associated costs have dropped dramatically over the past decade^17^. However, few studies have looked at the utility of WGS in CyKD on a large scale^18^.

Large-scale WGS datasets facilitate the assessment of the association of variants across the allelic spectrum with a certain phenotype in an unbiased genome-wide manner. This is especially useful in the study of the genomics of rare diseases, where approaches reliant on the sequencing of candidate gene panels or single genes of interest lead to discovery bias. In this study we perform genome-wide association analyses using WGS data from 1,209 CyKD cases and 26,096 ancestry matched controls. We perform variant-level, gene-level, pathway-level, and time-to-event association analyses including rare, common, and structural variants in a diverse-ancestry population providing the largest unbiased assessment of CyKD to date. We identify strong associations in known (*PKD1, PKD2, DNAJB11, IFT140)* and novel genes (*COL4A3,* monoallelic *PKHD1)* across various variant types. We leverage this data to conduct the largest common variant GWAS of CyKD to date, showing no significant associations, and use this to define the common variant heritability of CyKD as between 3-9%.

## Results

### The CyKD cohort demographics

1,558 participants were recruited to the 100,000 Genomes Project (100KGP) with cystic kidney disease. 1,294 were probands (full breakdown of recruited pedigrees in Supplementary Table 2a). The demographic information top five most frequent HPO codes of probands is shown in Table 1.

**Table 1.**
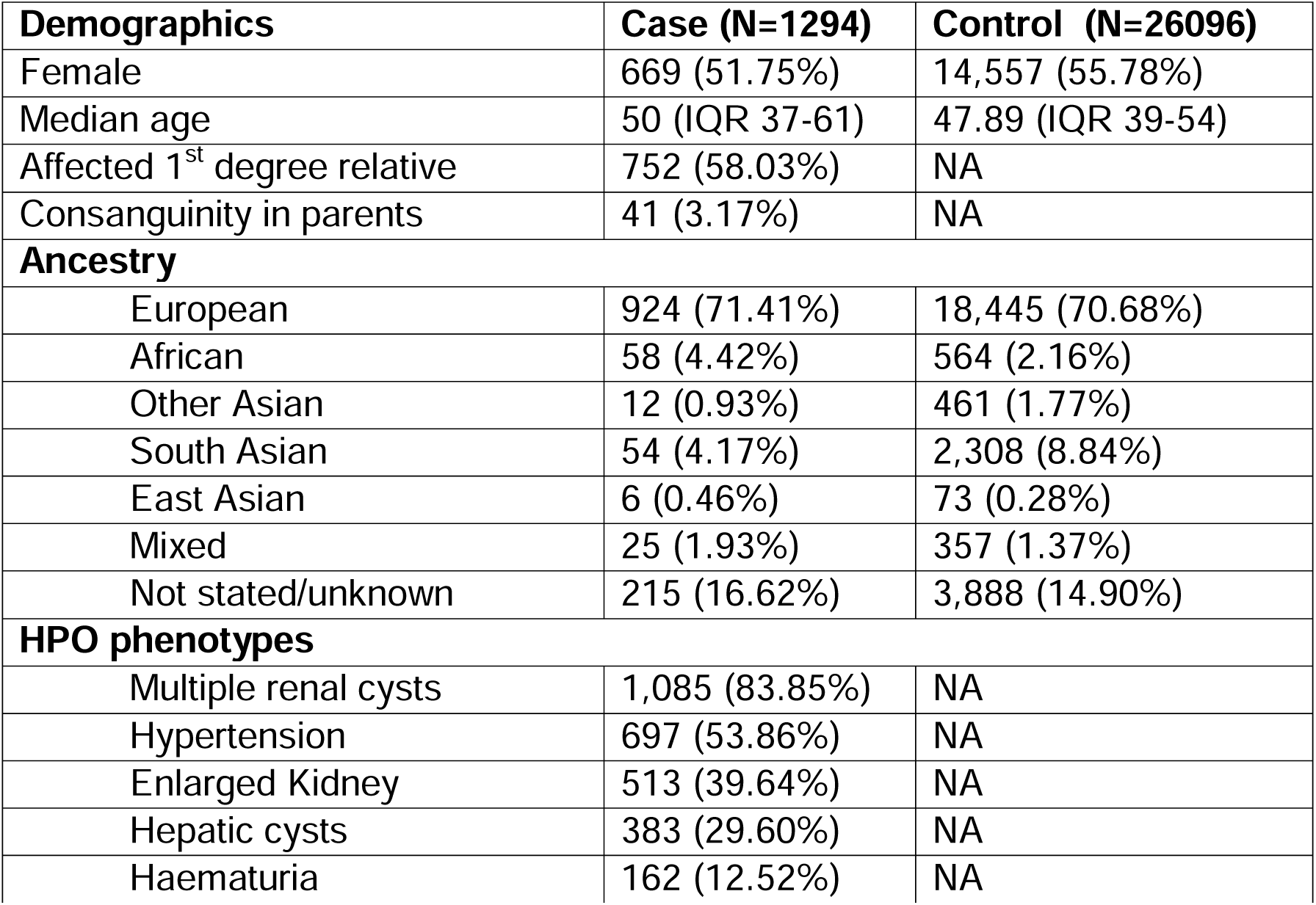
– Demographic and phenotypic breakdown of the recruited cystic kidney disease probands and controls.

| Demographics | Case (N=1294) | Control (N=26096) |
| --- | --- | --- |
| Female | 669 (51.75%) | 14,557 (55.78%) |
| Median age | 50 (IQR 37-61) | 47.89 (IQR 39-54) |
| Affected 1<sup>st</sup> degree relative | 752 (58.03%) | NA |
| Consanguinity in parents | 41 (3.17%) | NA |
| **Ancestry** |  |  |
| European | 924 (71.41%) | 18,445 (70.68%) |
| African | 58 (4.42%) | 564 (2.16%) |
| Other Asian | 12 (0.93%) | 461 (1.77%) |
| South Asian | 54 (4.17%) | 2,308 (8.84%) |
| East Asian | 6 (0.46%) | 73 (0.28%) |
| Mixed | 25 (1.93%) | 357 (1.37%) |
| Not stated/unknown | 215 (16.62%) | 3,888 (14.90%) |
| **HPO phenotypes** |  |  |
| Multiple renal cysts | 1,085 (83.85%) | NA |
| Hypertension | 697 (53.86%) | NA |
| Enlarged Kidney | 513 (39.64%) | NA |
| Hepatic cysts | 383 (29.60%) | NA |
| Haematuria | 162 (12.52%) | NA |

### The clinically validated arm of the 100KGP gave a molecular diagnosis to 53% of the CyKD cohort

Of these probands, 1,290 had a genetic diagnosis from the 100KGP clinical pipeline: 640 (52.93%) were solved, 34 (2.81%) were partially solved, 79 (6.54%) had missing data and 537 (44.42%) were unsolved. The full breakdown of solved cases and the types of variants can be found in Table 2 (three patients were solved for primary conditions unrelated to their cystic kidney disease e.g. intellectual disability and were not included in this table and 12 cases did not have a gene recorded despite being listed as solved).

**Table 2.**
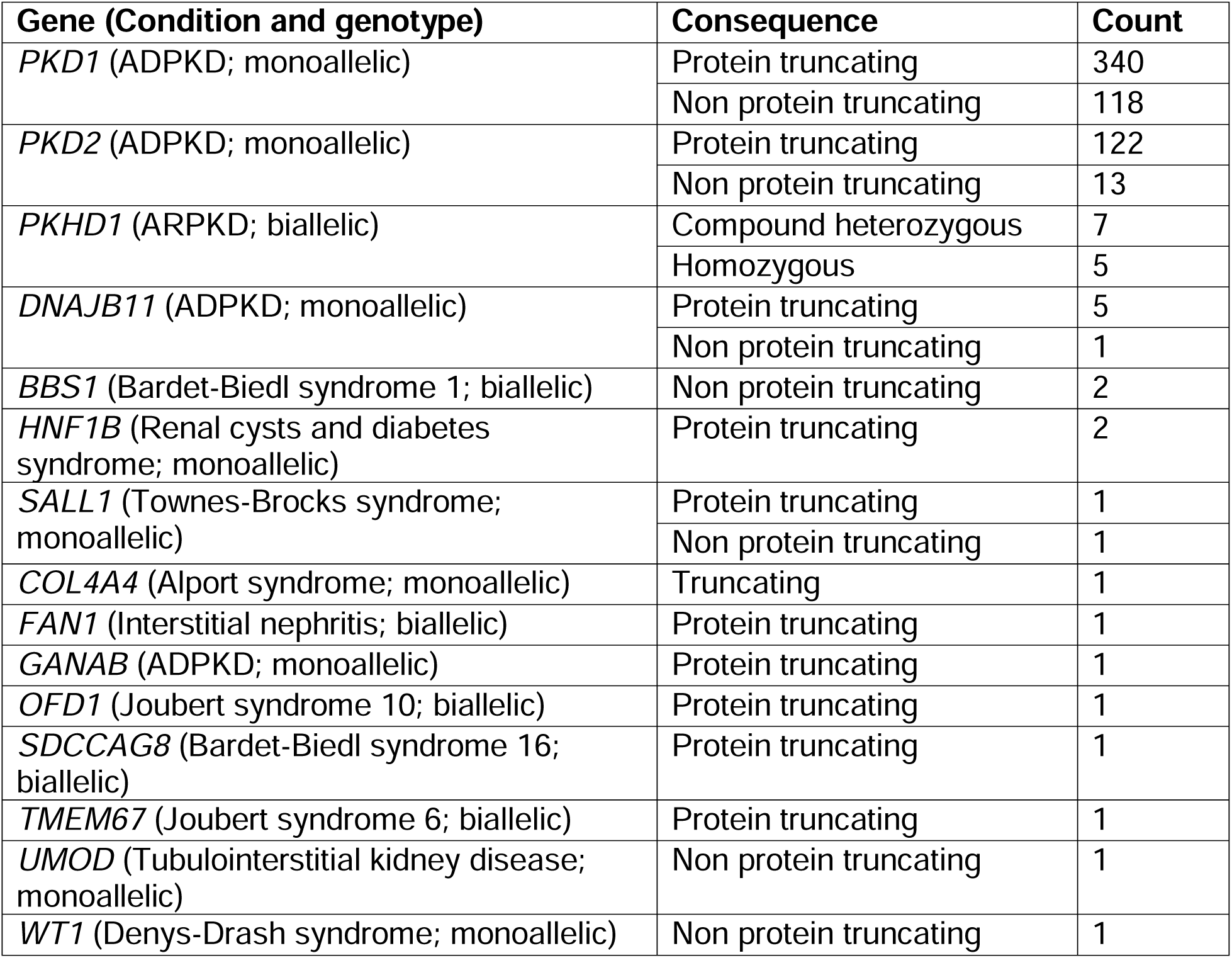
– Molecular diagnosis in cystic kidney disease cases that were solved by the 100,000-genomes project clinical pipeline.

| Gene (Condition and genotype) | Consequence | Count |
| --- | --- | --- |
| *PKD1* (ADPKD; monoallelic) | Protein truncating | 340 |
|  | Non protein truncating | 118 |
| *PKD2* (ADPKD; monoallelic) | Protein truncating | 122 |
|  | Non protein truncating | 13 |
| *PKHD1* (ARPKD; biallelic) | Compound heterozygous | 7 |
|  | Homozygous | 5 |
| *DNAJB11* (ADPKD; monoallelic) | Protein truncating | 5 |
|  | Non protein truncating | 1 |
| *BBS1* (Bardet-Biedl syndrome 1; biallelic) | Non protein truncating | 2 |
| *HNF1B* (Renal cysts and diabetes syndrome; monoallelic) | Protein truncating | 2 |
| *SALL1* (Townes-Brocks syndrome; monoallelic) | Protein truncating | 1 |
|  | Non protein truncating | 1 |
| *COL4A4* (Alport syndrome; monoallelic) | Truncating | 1 |
| *FAN1* (Interstitial nephritis; biallelic) | Protein truncating | 1 |
| *GANAB* (ADPKD; monoallelic) | Protein truncating | 1 |
| *OFD1* (Joubert syndrome 10; biallelic) | Protein truncating | 1 |
| *SDCCAG8* (Bardet-Biedl syndrome 16; biallelic) | Protein truncating | 1 |
| *TMEM67* (Joubert syndrome 6; biallelic) | Protein truncating | 1 |
| *UMOD* (Tubulointerstitial kidney disease; monoallelic) | Non protein truncating | 1 |
| *WT1* (Denys-Drash syndrome; monoallelic) | Non protein truncating | 1 |

### Outcome data shows those with pathogenic *PKD1* variants have the worst renal prognosis followed by those without a diagnosis

Of the 1,290 cases, 578 (44.8%) had data regarding kidney function in the form of HPO or HES codes of whom 398 (68.9%) had reached KF (full breakdown in Supplementary Tables 2b and S2c). Survival analysis of these cohorts followed what is known about the renal prognosis in variants in CyKD (Figure 2).

**Figure 1.**
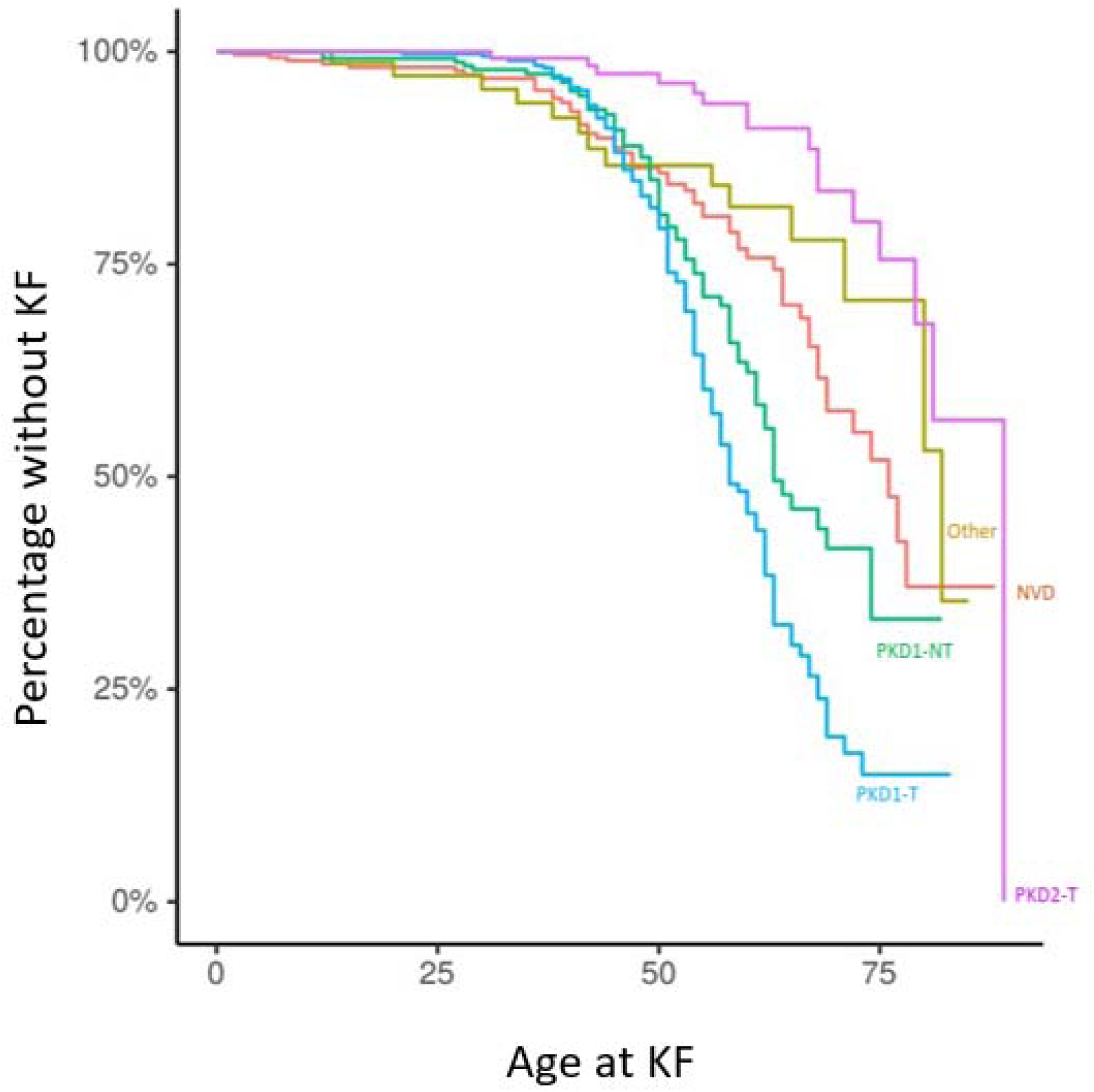
– Kaplan-Meier plot of kidney survival plotted by primary driving variant. PKD1-T PKD1-truncating variant, PKD1-NT PKD1-nontruncating variant, PKD2-T PKD2-truncating variant, Other-another variant in the PanelApp cystic kidney disease gene panel

**Figure 2.**
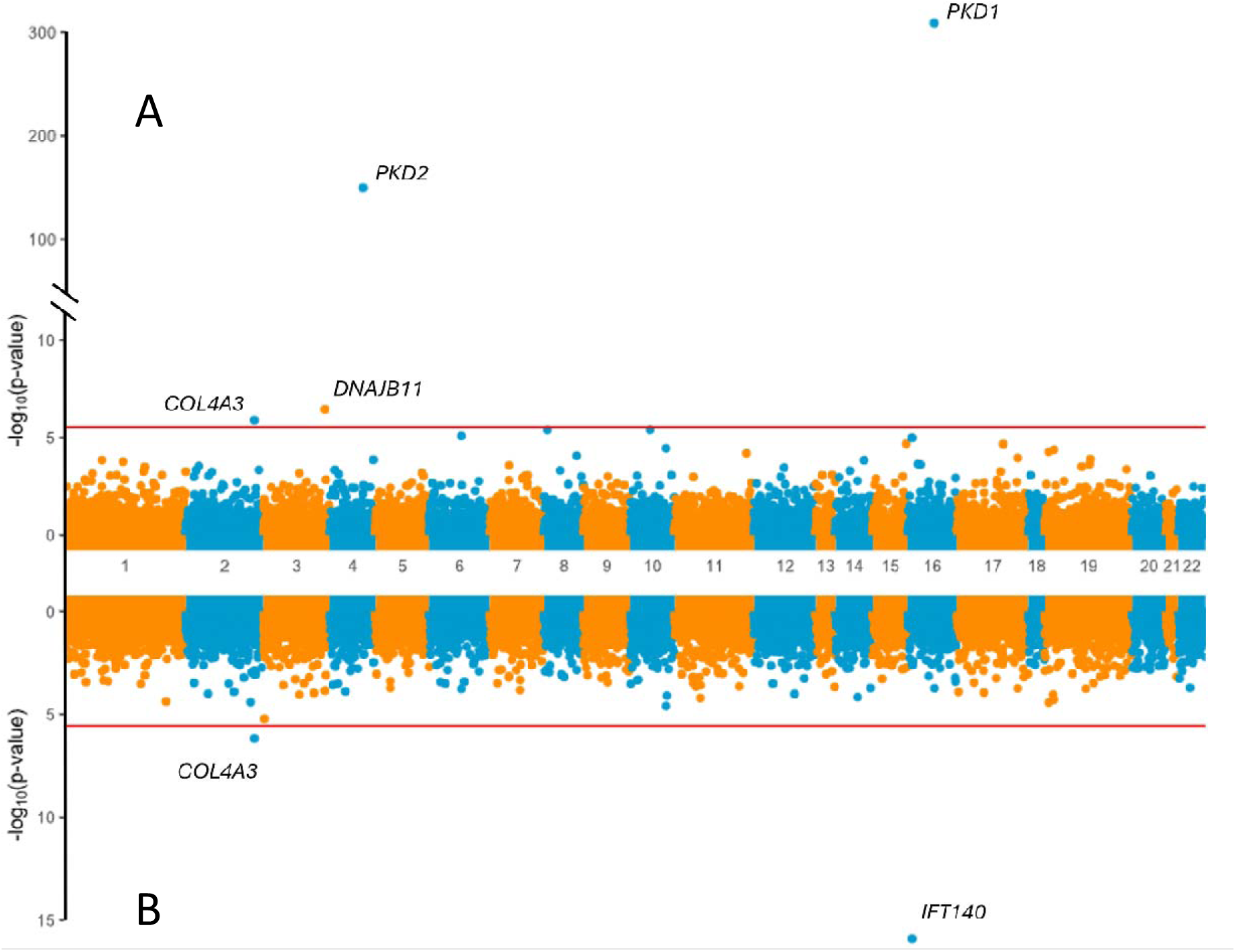
– Rare variant collapsing analysis under the missense+ mask. Gene-based Miami plots of the SAIGE-GENE “missense+” analyses. Each point is a gene, made up of variants that qualified under their respective mask. A. Missense+ analysis of the total ancestry matched cohort of 1,209 cases and 26,096 showing a significant enrichment of cases for PKD1, PKD2, DNAJB11 and COL4A3. B. Missense+ analysis of the depleted cohort of 308 cases versus 26,096 controls showing enrichment of cases for IFT140 and COL4A3. The horizontal line indicates the threshold for exome-wide significance.

### Combining the clinical pipeline with research grade diagnostics led to a genetic diagnosis for the majority of the ancestry matched CyKD cases

994 of the 1,209 (82%) tested cystic kidney disease cases had a monogenic or single structural variant contribution for their disease identified when combining the clinical pipeline results with those patients who had variants identified via collapsing gene-based analyses (Table 3). These are discussed in more detail below. Of note the research grade diagnostics cohort is the ancestry matched cohort and hence a lower number than the total CyKD probands.

**Table 3.** – Cohort breakdown by variant type in the 100KGP CyKD cohort after clinical and research pipeline.

| Variant type | Cases (n=1209)<br>attributable (% of total cohort) |
| --- | --- |
| *PKD1* truncating SNV (including splice variant) | 411 (34%) |
| *PKD1* non-truncating SNV | 249 (21%) |
| *PKD2* truncating SNV (including splice variant) | 128 (11%) |
| *PKD2* non-truncating SNV | 27 (2%) |
| *PKD1/2* SV | 33 (3%) |
| 17q12 deletions | 7 (1%) |
| *IFT140* SNV | 27 (2%) |
| *COL4A3* SNV | 15 (1%) |
| Other genes from PanelApp SNV | 79 (6%) |
| Monoallelic *PKHD1* SNV | 18 (1%) |
| No rare variant contributor detected | 215 (18%) |

### Unbiased rare variant analysis highlights *IFT140* and *COL4A3* as important genes involved in CyKD

Rare variant analysis of the total ancestry matched cohort of 1,209 cases and 26,096 controls under the “missense+” mask showed a significant enrichment of cases for *PKD1 (P=1.17×10^−309^, OR=10.60, 95% CI = 9.35-12.01), PKD2 (P=1.96×10^−150,^ OR=19.07, 95% CI 15.13-23.99), DNAJB11 (P=3.52×10^−7^, OR 1.07, 95% CI 0.95-1.21), and COL4A3 (P=1.26×10^−6^, OR=3.02, 95% CI 2.10-4.22. See* Figure 3a). There was no evidence of genomic inflation (lambda<1, see QQ-plot in Supplementary Figure 2b).

**Figure 3.**
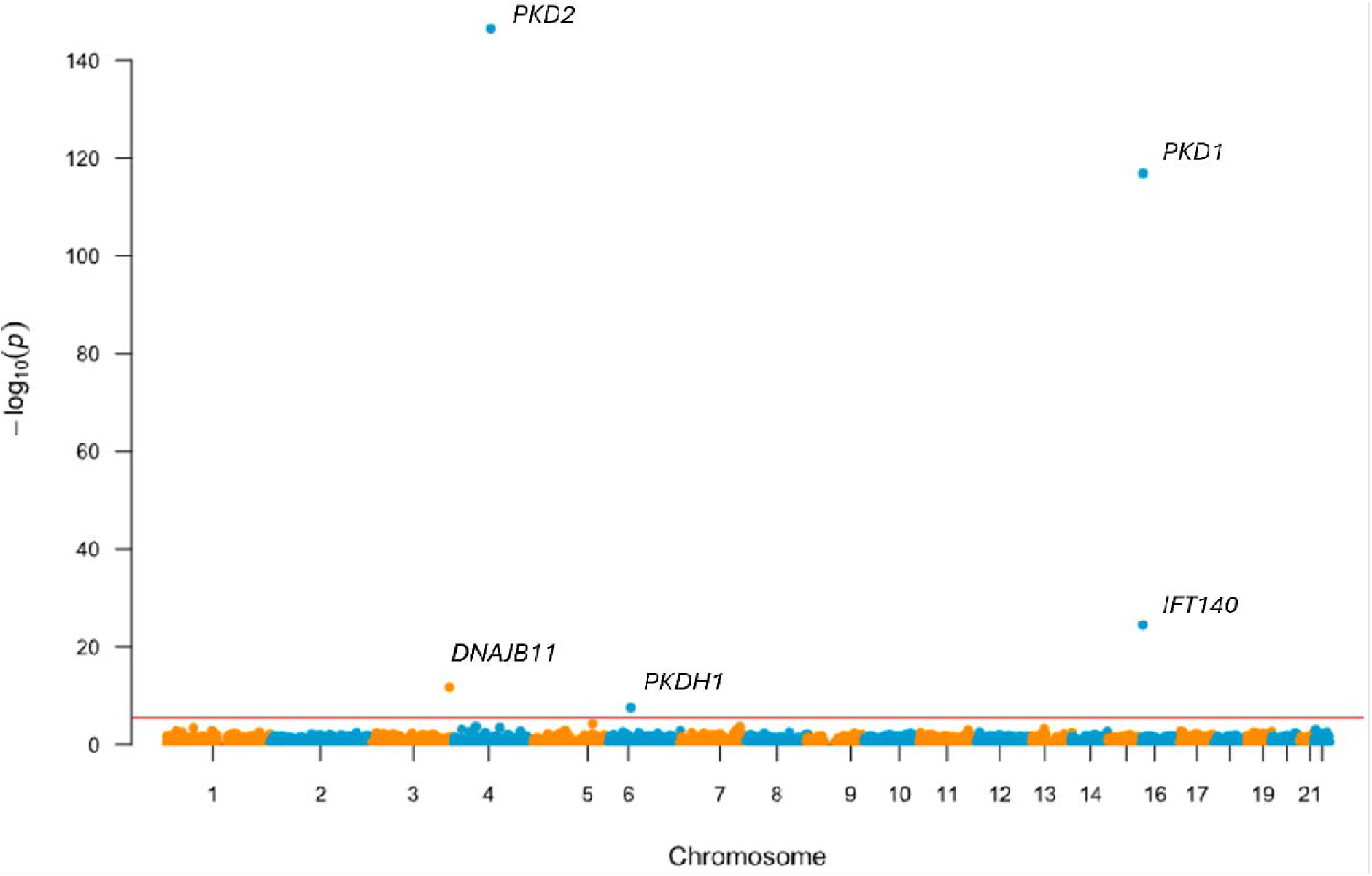
– Rare variant collapsing analysis under loss-of-function mask. Gene-based Manhattan plot of the SAIGE-GENE “loss-of-function” analysis of the total ancestry matched cohort of 1,209 cases and 26,096 showing a significant enrichment of cases for PKD1, PKD2, DNAJB11, IFT140 and PKHD1. Each point is a gene, made up of variants that qualified under their respective mask. The red line indicates an exome-wide significance level of P=2.5×10^−6^.

Removing cases solved by 100KGP and patients that had a bioinformatically ascertained pathogenic variant in a known cystic gene left 308 cases (at the time of analysis *IFT140* was not a known CyKD gene). Repeating the rare variant analysis under the “missense+” mask in this group showed a significant enrichment of cases with variants in *IFT140 (P=1.26×10^−16^, OR=5.57, 95% CI 3.63-8.21)* and *COL4A3 (P=6.83×10^−7^, OR=4.93 95% CI 2.77-8.11)* compared with 26,096 controls (Figure 3b).

### *IFT140* and *COL4A3* variants in unsolved individuals most likely represent their primary diagnosis

27 cases (8.8%) within the 308 unsolved cases had a qualifying variant in *IFT140* under the “missense+” mask. Of the 27 cases, all were heterozygous for the qualifying variants. None of the variants individually reached genome-wide significance.

Analysis of the SVs and CNVs within *IFT140* revealed two additional cases (0.65%) with heterozygous exon crossing SVs from the 308 unsolved cystic disease cases. There was one 3.6kb deletion spanning exon 16 and 17 and one patient with two different small (12.6kp and 8.1kb), exon crossing inversions. Four exon crossing SVs (three deletions and one tandem duplication) were seen in four different controls (0.015%). There was enrichment of *IFT140* SVs in cases versus controls (*P=* 0.0032) although this was not significant at a genome-wide level. None of the 27 initial cases with *IFT140* SNVs had detectable CNVs affecting *IFT140* compared to three CNVs seen in 26,096 controls (*P=* 1).

There were no plausible second variants within *IFT140* in any of these individuals. A full variant and phenotypic breakdown of the *IFT140* cases can be found in Supplementary Table 3a.

Amongst the 15 unsolved cystic kidney disease patents with qualifying variants in *COL4A3* under the “missense+” mask all were heterozygous for their respective variants and did not overlap with the unsolved *IFT140* cohort listed above. None of the variants individually reached genome-wide significance. A full variant and phenotypic breakdown of the *COL4A3* cases can be found in Supplementary Table 3b.

### Analysis of loss-of-function (protein truncating) variants identifies monoallelic defects of *PKHD1* in unsolved CyKD

Collapsing rare variants that had a high confidence call for loss-of-function under the “LoF” mask (i.e. analysis restricted to protein length-altering variants, excluding all missense variants) revealed significant enrichment of cases for *PKD2 (P=3.05×10^−147^, OR=130.85, 95% CI 83.66-215.37), PKD1 (P=1.29×10^−117^,OR=36.01, 95% CI 30.52-42.23), IFT140 (P=3.00×10^−25^, OR=14.03, 95%CI 7.91-24.45), DNAJB11 (P=1.84×10^−12^, OR 1.07, 95% CI 0.95-1.21) and PKHD1 (P=2.98×10^−08,^ OR=4.07 95%CI 2.24-6.88)* (Figure 4).

**Figure 4.**
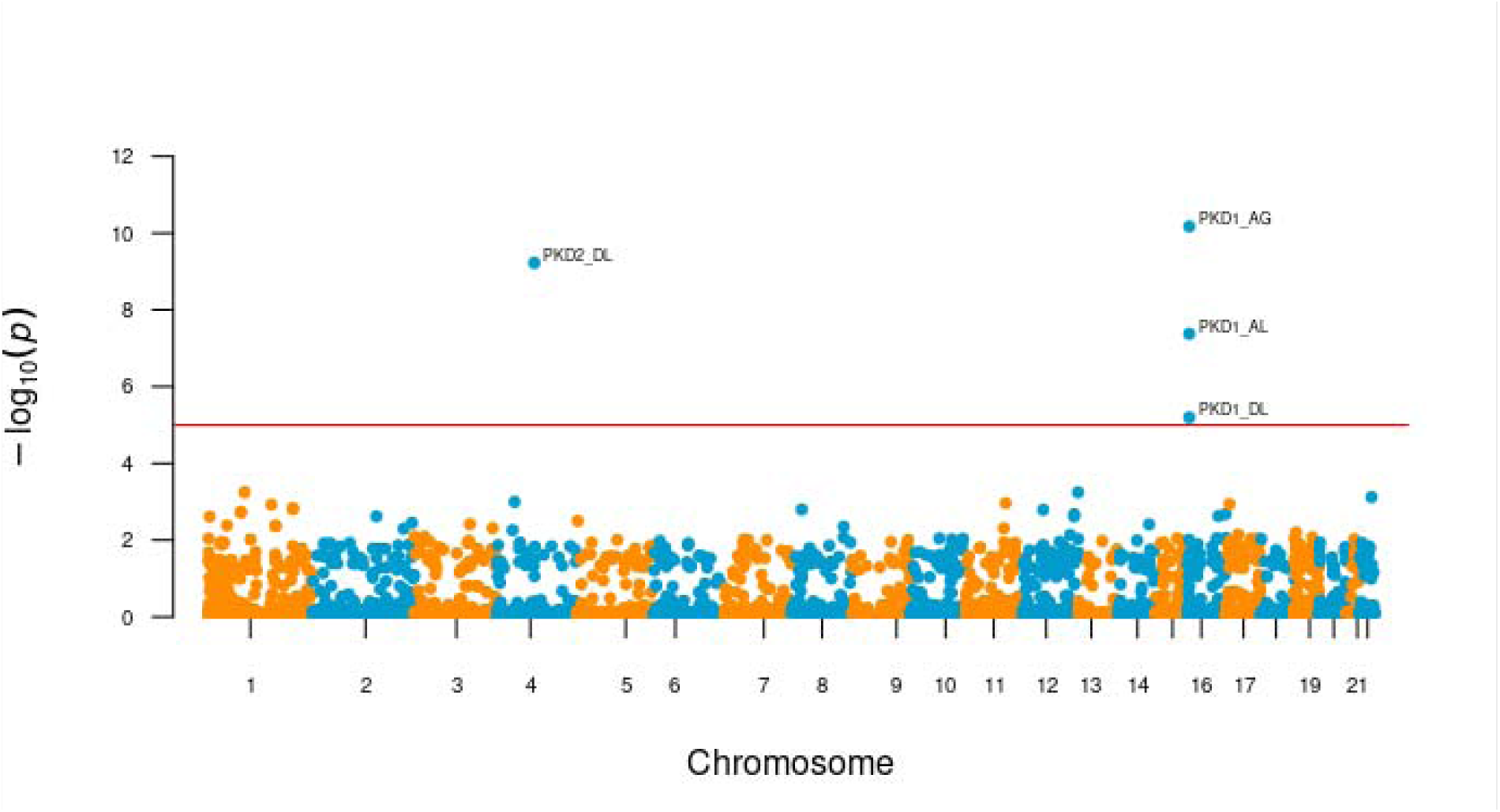
– Gene based Manhattan plot of the SAIGE-GENE analysis with the splice mask. Each point is a gene representing the significance of the association with cystic kidney disease in 248 cases versus 26096 controls, made up of variants that are highly likely (spliceAI score >0.8) to impact splicing. The horizontal line indicates the threshold for genome wide significance (genome or exome? Add the threshold used). There was significant enrichment in acceptor gain (AG), acceptor loss (AL) and donor loss (DL) variants for PKD1 (AG P= 6.70×10-11, AL P= 4.22×10-08, DL P=6.32×10^−6^) and for DL in PKD2 (P=5.97×10^−10^).

Further depletion of cases by removing those with qualifying variants that made up the *IFT140* and *COL4A3* signals led to 266 cases remaining which did not reveal any further significant associations on rare variant testing (Supplementary Figure 2).

In all presented analyses, the patients were heterozygous for their qualifying variants except in *DNAJB11* where 59 of the 369 cases that had qualifying variants within the “missense+” mask were homozygous.

There were 61 predicted LoF variants in *PKHD1* that made up the association signal in the LoF mask analysis of the whole cystic disease cohort. These were seen in 50 cases of which 22 were solved, two were partially solved, 24 were unsolved and two were unascertainable. All 50 cases were heterozygous for the variant that made up the signal.

Of the 22 solved cases, three patients had a diagnosis of ARPKD secondary to biallelic *PKHD1* variants, and 19 had a diagnosis of ADPKD due to variants in *PKD1* or *PKD2*. In the two partially solved cases both patients had a second *PKHD1* variant deemed to be a variant of unknown significance (VUS).

Of the 24 unsolved cases with a single LoF *PKHD1* variant, four also had a computationally predicted high impact non-truncating variant in *PKD1*, and one (in addition to the *PKHD1* variant) had a predicted high impact non-truncating *PKD2* variant.

In the remaining 18 cases with a single heterozygous *PKHD1* LoF variant there were no SNVs, SVs or CNVs that would imply compound heterozygosity (and a diagnosis of ARPKD), or potentially pathogenic variants in any other gene associated with CyKD. Two patients had a second *PKHD1* variant with CADD >20 in *PKHD1,* but both had been deemed “likely benign” by Clinvar (Clinvar ID: 1187104 and 102305).

In total, 634 (2.4%) of the 26,096 controls carried qualifying monoallelic *PKHD1* LoF variants. When compared to the 18 (6.7%) out of 266 unsolved cases with no clear molecular diagnosis there is a significant enrichment of *PKHD1* variants in the unexplained cystic disease cohort (P=5.85×10^−6^, OR=2.92, 95% CI 1.69-4.76). Three of the 18 monoallelic *PKHD1* cases had reached KF at a median age of 42 years. There was no statistical difference between the rates of liver cysts between monoallelic *PKHD1* cohort and the general CyKD cohort (*P=0.31*). The full demographic details of the *PKHD1* cohort can be found in Supplementary table 3c.

### Non-coding collapsing analysis of the no variant detected (NVD) cohort revealed enrichment of splice site variants in PKD1 and PKD2

Removing the cases with qualifying *IFT140*, *COL4A3* and monoallelic *PKHD1* variants led to no further enrichment in the NVD cohort under the “missense+” or “LoF” gene collapsing tests. However, in the remaining 248 cases versus 26,096 controls there was significant enrichment in acceptor gain (AG), acceptor loss (AL) and donor loss (DL) splice variants for *PKD1 (AG P= 6.70×10^−11^ OR=150.57 95% CI 35.39-730.24, AL P= 4.22×10^−8^ OR=398.51 95% CI 39.10-16384, DL P=6.32×10^−6^ OR=no variants in controls)* and for DL in *PKD2 (P=5.97×10^−10^ OR=no variants in controls)* (Figure 5).

**Figure 5.**
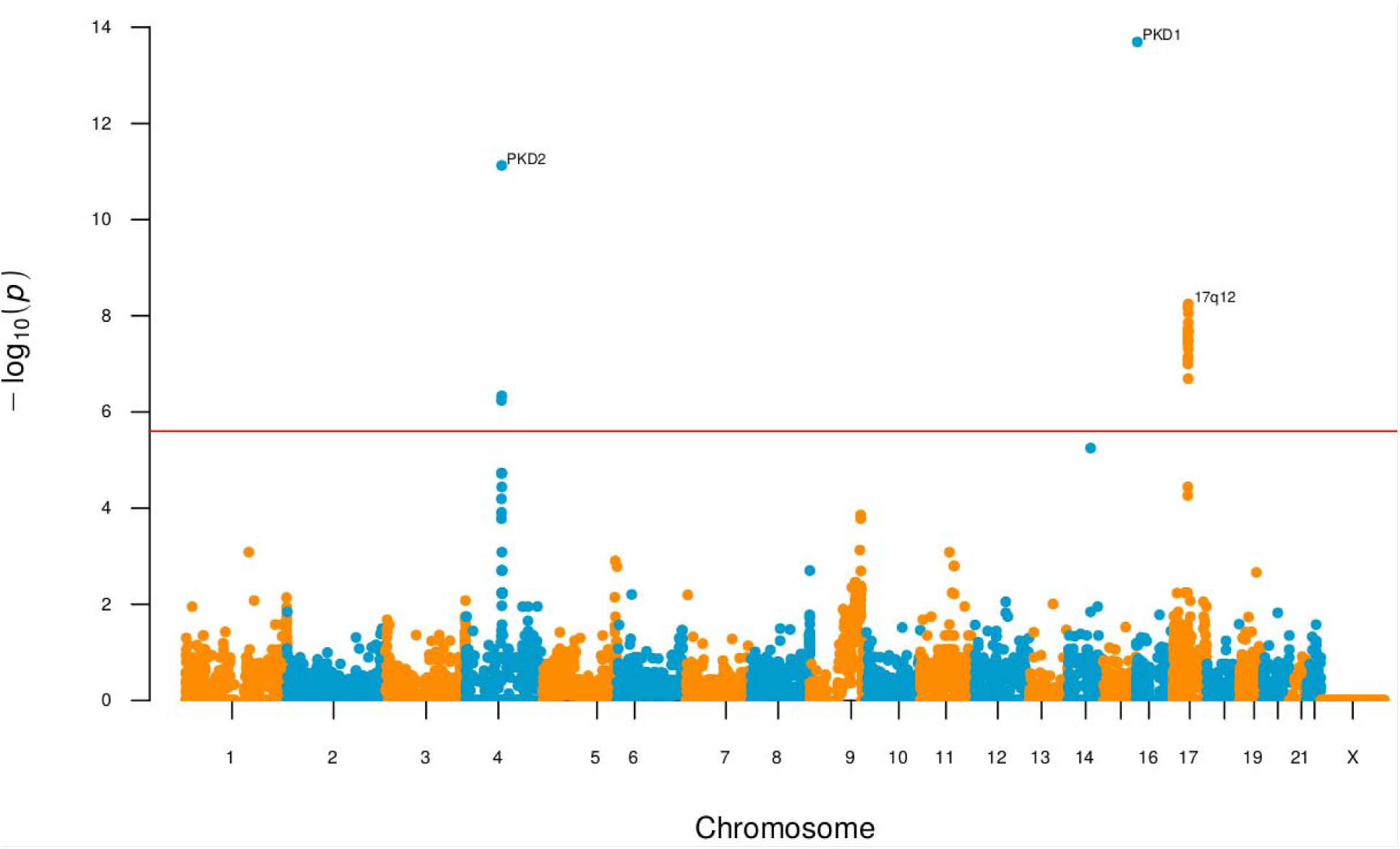
– Gene based Manhattan plot of the exome-wide gene based structural variant analysis. Each point is a gene representing the significance of the association with cystic kidney disease in 1,209 cases and 26,096 controls, made up of rare (in-house MAF<0.01), exon crossing SV/CNVs that have been called by MANTA/CANVAS. Common SV/CNVs (MAF>0.1%) seen in gnomAD or the 100KGP cancer cohort were excluded. The horizontal line indicates the threshold for genome-wide significance. The significant associations were PKD1 (P=2.02×10^−14^), PKD2(P=7.48×10^−12^) and the 17q12 locus (P=8.81×10^−9^).

There was no further enrichment in the 3’ or 5’-UTR regions, intronic regions with a CADD score >20 or donor gain splice sites on a genome-wide basis.

### Rare variant analysis by primary variant does not reveal contribution of variants in other genes

Using the primary variant, the cystic cohort was divided into those cases with *PKD1* and *PKD2* truncating and non-truncating variants, respectively. Bar the primary gene in each cohort there was no further significant enrichment of any other gene.

A full list of the summary statistics which includes the variants that make up each association can be found in Supplementary Tables 4-19.

### Structural variants in *PKD1, PKD2* and the 17q.12 loci play an important role in cystic kidney disease

Exome wide gene-based SV analysis was performed in all cystic kidney cases and ancestry matched controls. Across all combined types of SV there was significant enrichment in *PKD1 (P=2.02×10^−14^, OR=2.52 95% CI 1.69-3.63), PKD2 (P=7.48×10^−12^, OR=3.51, 95% CI 1.74-6.37)* and genes within 17q12 locus including *HNF1B (P=8.81×10^−9^, OR=7.11, 95% CI 3.41-13.66).* Of note, two genes within proximity of *PKD2* also reached genome-wide significance: *SPARCL1 (P=5.76×10^−7^)* and *HSD17B11 (P=8.69×10^−6^)* but these were made up of large CNVs that also encompassed *PKD2* (Figure 6).

**Figure 6.**
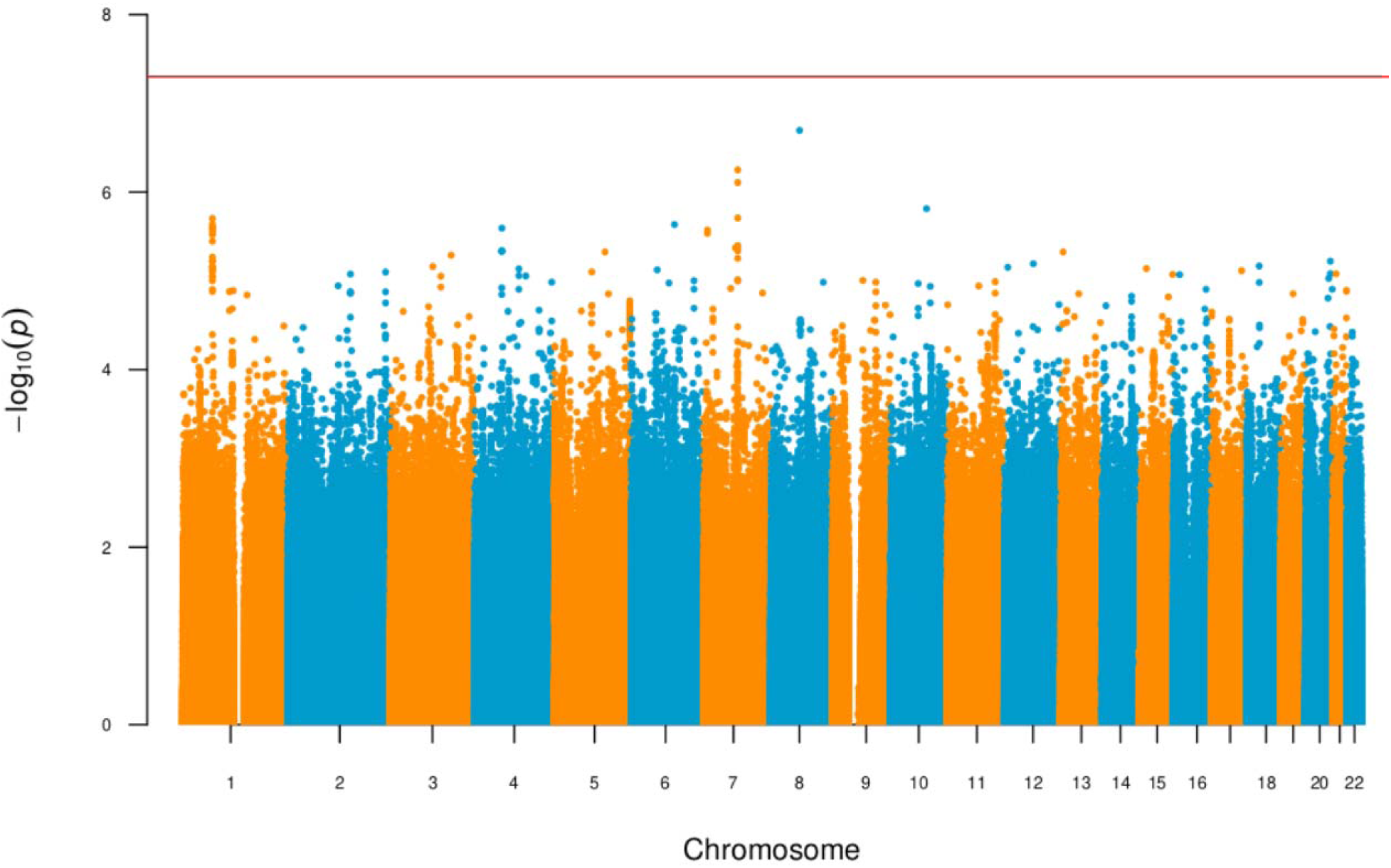
– Manhattan plot of CyKD metanalysis. Manhattan plot of CyKD metanalysis across 6,641,351 markers in 2,923 cases (1,209 GEL, 780 FinnGen, 510 JBB, 424 UKBB) and 900,824 controls (26,096 GEL, 341,081 FinnGen, 178,216 JBB, 355,431 UKBB). There is no variant that reaches genome-wide significance. The genomic inflation is 1.012.

The *PKD1* signal was predominantly driven by small deletions <10kb (median size 1.14kb, IQR 0.46-3.06kb) (*P=2.17×10^−22^, OR=8.11 95% CI 4.58-13.83).* For *PKD2* (*P=7.48×10^−12,^ OR=13.03 95% CI 5.02-31.87*) and the 17q12 locus *(P=4.12×10^−8^, OR=8.70, 95% CI 3.72-18.80),* the signal was driven by deletions >10kb (median size in *PKD2* 405kb, IGR 98-1371kb; 17q12 1550kb, IQR 1545-1639kb) with no other loci reaching genome-wide significance. No genes reached genome wide significance for duplications.

Of the 46 patients with rare exon crossing SVs in *PKD1* or *PKD2,* 13 also harbored predicted LoF variants in *PKD1* or *PKD2,* thus leaving 33 patients with cystic kidney disease attributable to SVs in *PKD1* or *PKD2*.

Of the 11 patients with 17q12 loci CNVs in the cystic disease cohort, one patient had a *PKD1* non-truncating SNV and two had *PKD1* truncating SNVs that met the criteria for being likely causative. One patient had a known *HNF1B* CNV detected by a separate diagnostic lab prior to the return of 100KGP results.

Analyzing the subgroup of patients without an identified molecular diagnosis (n=248), there was significant enrichment for large (>10kb) deletions at the 17q12 loci (*P=9.21×10^−9^, OR=24.04 94% CI 8.00-60.71)* which were detected in seven probands.

Of the seven 17q12 patients, the median age was 13.5 years, significantly lower than the total cystic disease cohort (*P<0.05).* None of the patients had reached ESKD or had HPO or HES codes pertaining to diabetes; a full breakdown of phenotypic profile can be found in Supplementary Table 20 and all summary statistics from the SV analysis can be found in Supplementary Table 21-22.

### More recently described genes in CyKD are less penetrant than *PKD1* and *PKD2*

There was marked variation in the proportion of individuals with each gene/variant type that were documented to have cystic kidney disease, with the figures broadly comparable between 100KGP and UKBB (Table 4).

**Table 4.**
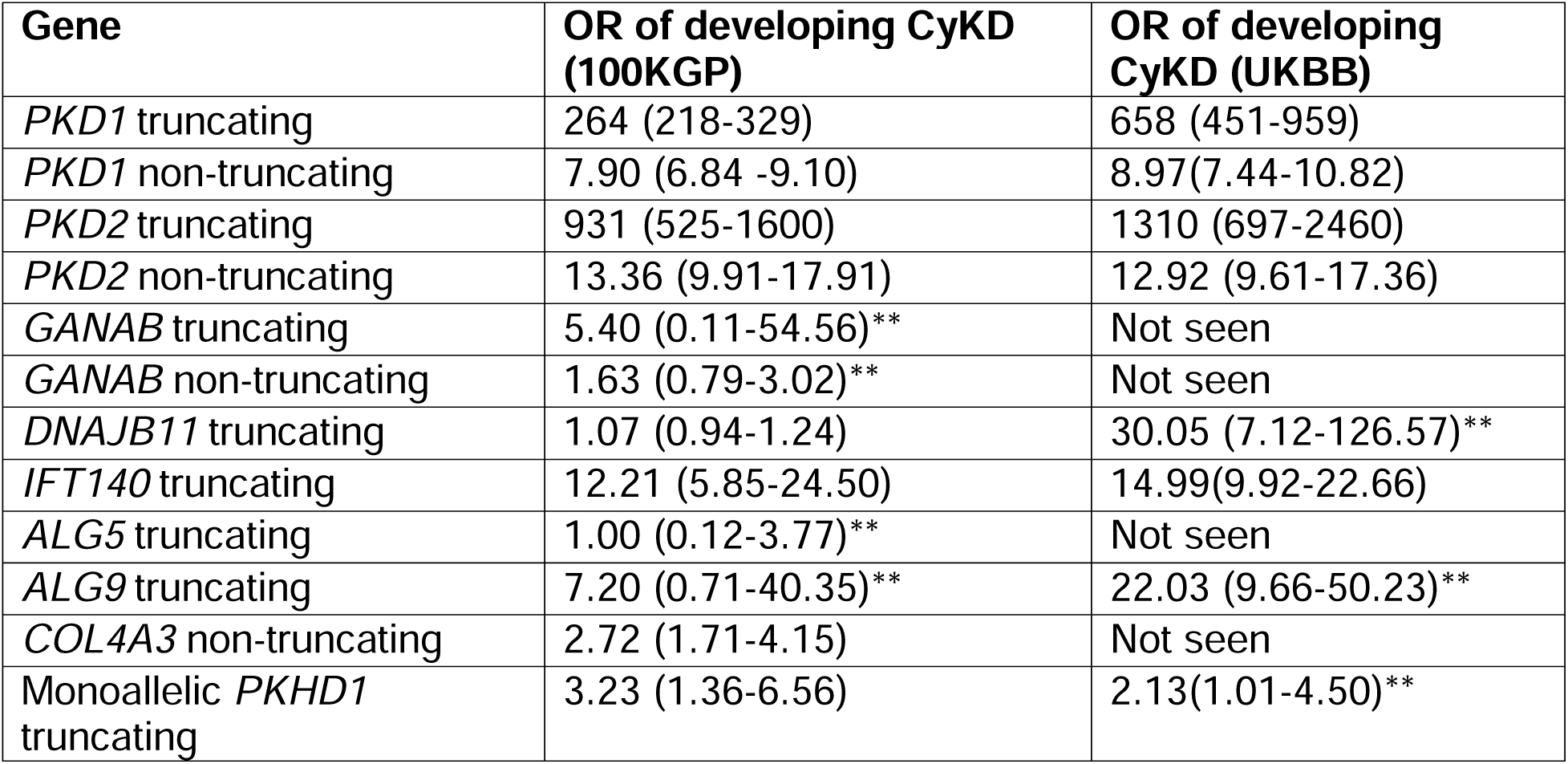
– Odds ratio of developing CyKD in the 100KGP (n=741) and the UKBB (n=825) in each different gene. OR for UKBB taken from the AstraZeneca analysis using the model closest to our analysis. **Association not statistically significant in association analysis, included here as they are clinically reportable genes for this phenotype. The 100KGP results are presented for those individuals who were between 40-70 years old at the time of recruitment to match the UKBB recruitment analysis.

| Gene | OR of developing CyKD (100KGP) | OR of developing CyKD (UKBB) |
| --- | --- | --- |
| *PKD1* truncating | 264 (218-329) | 658 (451-959) |
| *PKD1* non-truncating | 7.90 (6.84 -9.10) | 8.97(7.44-10.82) |
| *PKD2* truncating | 931 (525-1600) | 1310 (697-2460) |
| *PKD2* non-truncating | 13.36 (9.91-17.91) | 12.92 (9.61-17.36) |
| *GANAB* truncating | 5.40 (0.11-54.56)** | Not seen |
| *GANAB* non-truncating | 1.63 (0.79-3.02)** | Not seen |
| *DNAJB11* truncating | 1.07 (0.94-1.24) | 30.05 (7.12-126.57)** |
| *IFT140* truncating | 12.21 (5.85-24.50) | 14.99(9.92-22.66) |
| *ALG5* truncating | 1.00 (0.12-3.77)** | Not seen |
| *ALG9* truncating | 7.20 (0.71-40.35)** | 22.03 (9.66-50.23)** |
| *COL4A3* non-truncating | 2.72 (1.71-4.15) | Not seen |
| Monoallelic *PKHD1* truncating | 3.23 (1.36-6.56) | 2.13(1.01-4.50)** |

### SeqGWAS of CyKD reveals no replicable common variant associations

A seqGWAS of 1,209 CyKD cases and 26,096 ancestry-matched controls using 10,377,275 variants with a MAF>1% (Supplementary Figure 3) revealed only a single variant reaching genome-wide statistical significance on chromosome 8, chr8:92259567:A:C (*P=1.38×10^−8^, OR 0.72).* There was no evidence of genomic inflation (lambda 0.99). To confirm/refute this surprising finding we meta-analyzed this dataset with those from the UK, Japanese and FinnGen biobanks. In the non-100KGP datasets there was evidence of association at several loci, most notably a stop gain in *PKHD1* in the FinnGen cohort (see Supplementary Figures 4-5 for individual study Manhattan plots) but the chr8:92259567:A:C signal was not replicated so is inferred to be a false positive, and overall in the combined analysis of 2,923 cases and 900,824 controls across 6,641,351 markers there were no genome-wide significant variants (Figure 7).

**Figure 7.**
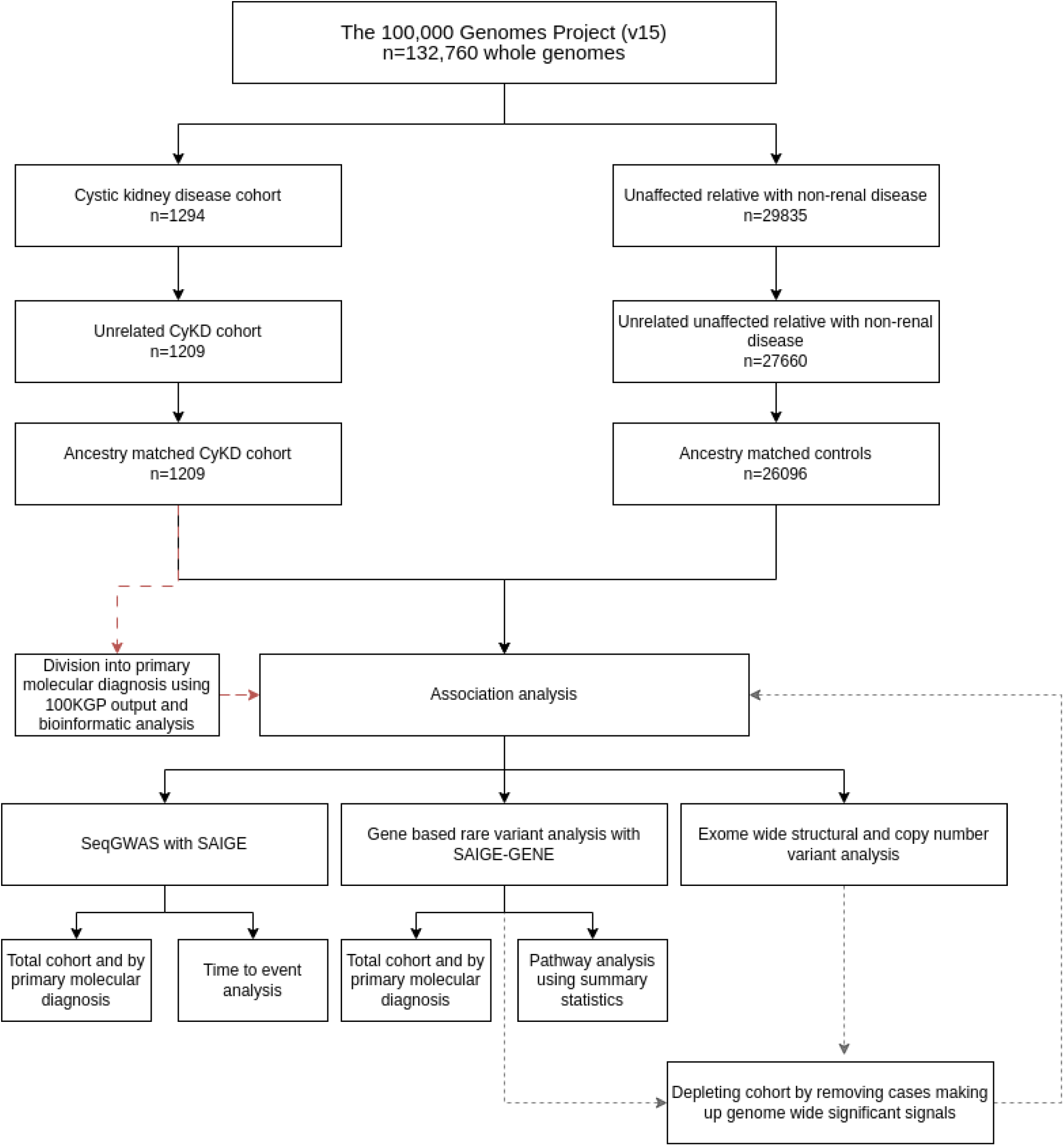
– Study workflow highlighting the creation of the cohort and analysis steps. Cases and controls underwent relatedness analysis and ancestry matching before inclusion in the association analyses. The cases were also subdivided into their primary driving variant and association testing performed as subgroup analyses. For the main body of the analysis whenever genome wide significant genes were discovered, the cases making up the signal were removed from the cohort and the analysis repeated until no further significant signal was detected.

Subgroup analysis by primary disease-causing variant type did not reveal any genome-wide significant loci (see Supplementary Figure 6 and 7).

### The proportion of heritability attributable to common variants was between 3%-9%

Within 100KGP the proportion of phenotypic variance (h) explained by additive common and low-frequency variation among individuals of European ancestry with CyKD was 9.0% (SE 7.6%). Using the summary statistics from the combined FinnGen/UKBB CyKD GWAS the estimated heritability was 3.0% (SE 9.7%). The large standard errors reflect low power to detect heritability within this cohort.

### Time to event analysis did not reveal any trans-acting genetic modifiers of severity

Within the cohort 398 of the 1,288 probands had reached KF (30.9%) with a median age of 52 years (IQR 44-60). Time to event analysis using GATE did not reveal any genome-wide significant associations – either in the total cohort or stratified by primary gene or variant type (see Supplementary figure 8).

## Discussion

994 of 1,209 (82%) cystic kidney disease patients had a monogenic SNV or SV cause for their disease identified through a combined clinical grade and unbiased research analyses of biobank-scale WGS data.

This high diagnostic yield of WGS in to investigate CyKD practice has led to this technology being made available to all suspected CyKD patients in the UK via the National Health Service’s Genomic Medicine Service^45^ (though it must be noted that as yet a proportion of these variants do not necessarily meet ACMG criteria for issuing a clinically actionable molecular diagnosis).

The data presented also clarifies the underlying genetic architecture of cystic kidney disease. They point strongly to the conclusion that cystic kidney disease is extensively driven by monogenic mechanisms via rare variants of multiple different types, with a small contribution from common variants.

The arguments for this position are compelling. Firstly, this unbiased method has confirmed the importance of established and newly described genes in the pathogenesis of cystic kidney disease (*PKD1*, *PKD2*, *IFT140, DNAJB11*).

Secondly, we provide robust statistical evidence that *COL4A3 is* associated with cystic kidney disease. Smaller studies have hinted at this association^46^ and sequencing of unexplained KF patients in an American cohort, showed a significant proportion of unexplained cystic cases were attributed to the *COL4A* family of genes^47^.

Thirdly, our attempts to find more complex “common” variants via seqGWAS that contribute to the CyKD phenotype by meta-analyzing more than 2,000 cases with nearly one million controls did not reveal any significant associations. In fact, in the Finnish population that has undergone significant genetic bottlenecks causing positive selection for certain recessive variants there is an enrichment of a known pathogenic *PKHD1* variant at an allelic frequency that borders the “rare” variant mask (rs137852949, MAF in Finnish population = 7.48×10^−3^, MAF in non-Finnish European population = 3.24×10^−4^) but meets inclusion in FinnGen. This variant has been implicated as a heterozygous cause of polycystic liver disease^48^ and its enrichment in our cohort as a heterozygous entity provides evidence for its role as a monoallelic cause of cystic kidney disease.

Our analysis of common-variant heritability does suggest that 3%-9% of disease may be explained by low penetrance common variation though our efforts to ascertain this were underpowered resulting in large standard errors. The small magnitude of this contribution explains why individual genome-wide significant variants were not detected – the small effect size means an additional 440 cases would be required to reach power to detect a heritability signal with >80% confidence and that substantially larger studies would be needed to detect the loci contributing to this risk. (A full power calculation can be found in supplementary figure 9). Our efforts to use age of KF per driving molecular diagnosis within CyKD is an attempt to try and unpick common variant contributions to disease severity and quantitatively define genetic modifiers, a key question within CyKD, but power was not sufficient to detect any significant signals. This represents the largest systematic analysis of whether oligogenic or polygenic mechanisms are important in the etiology of CyKD and with a rapidly increasing number of CyKD patients undergoing WGS as part of routine clinical care in the UK and establishment of a National Genomic Research Library, sets up an analytical pathway to address this in the future with ever-increasing power.

Similar to our recent work on urinary stone disease highlighting the importance of intermediate effect rare variants as a risk factor^49^, as well as work describing an uncommon UMOD variant (present in 0.1% of European ancestry individuals) that confers an intermediate level of risk of KF^50^, we show that CyKD represents another disease enriched for such variation. As shown in Table 5, the odds ratio for CyKD diagnosis in each gene across both this cohort and the UK Biobank is highly variable, highlighting that pathogenic variants in some of the more recently reported genes associated with CyKD are likely to have such a low penetrance that they may seldom exhibit Mendelian patterns of inheritance in families and may be perhaps regarded clinically as intermediate risk factors for developing CyKD. Communicating this information clearly to patients and their relatives is likely to be important when counselling them about the pros and cons of predictive testing for these disorders.

Finally, our findings are replicated in the UK Biobank with the top gene associations with cystic kidney disease being *PKD1 (P=9.83×10^−^*^63^*), PKD2 (P=1.64×10^−60^)* and *IFT140 (P=4.52×10^−15^)* in a cohort of 531 patients and 239,516 controls^51^. We calculate that the UK Biobank CyKD cohort is powered to detect genes that explain ≥8% of the total phenotypic variance, meaning genes associated with smaller effect sizes are unlikely to be detected.

Using WGS we have also undertaken the first systematic assessment of the structural and non-coding variant contribution to CyKD. These contribute to unsolved cases highlighting the power WGS has in identifying sites previously untested by traditional sequencing techniques. Whilst splice site variants have been implicated in individual families with unexplained CyKD^52,53^ this analysis gives quantitative statistical evidence at a population level that suggests these sites should be scrutinized in clinical analysis of *PKD1* and *PKD2*, something that is increasingly being recognized^54^. Equally, utilizing methodology similar to the gnomAD SV working group^38^ we find, as they did, that SVs play a larger role in the variance landscape than previously thought. Whilst SVs in cystic kidney genes have been implicated in small numbers, this cohort level analysis attributes at least 3.35% (40/1209) of the cystic disease burden to SVs, a similar proportion to the recently described *IFT140* gene at a population level. These discoveries are made possible by WGS which, unlike older methods, such as multiplex ligation-dependent probe amplification (MLPA) or arrays, enable accurate calling of many different types of SVs genome-wide. These findings should help inform decisions about the sensitivity of short-read WGS and other potential sequencing approaches in the clinical setting such as RNA-sequencing or long read DNA^55^ sequencing.

In the remaining unsolved cases we did not find any further enrichment at a variant, gene, pathway or SV level. Given the findings above, one explanation is that we lacked the power to detect additional monogenic signals in this group – either because they have reduced effect size or are individually extremely rare. Alternatively, it may be that a proportion of this group exhibited cystic kidney disease as a consequence of non-monogenic developmental disorders or undocumented environmental exposures, such as to lithium^56^. Irrespective, this work gives an estimate of the cohort size (an additional 2000 cases using the assumptions in the power section of the methods) needed to power future studies to discover additional monogenic causes of CyKD using unbiased genome-wide approaches. As more patients with CyKD are sequenced as part of their routine healthcare in the UK it is possible that this threshold will be passed, and further monogenic causes will be discovered using this type of methodology. Coupled with developments in analytical techniques, identification of variants across the allelic frequency and disease risk spectrum may further extend understanding of the biological basis of cystic kidney disease.

This study has several notable limitations. From a phenotype perspective we are reliant on KF as our only marker of outcome and were unable to access granular phenotypic and imaging data for our cohort, limiting our ability to apply several prognostic tools^11,57^ that would have aided in further stratification. Secondly, whilst this study represents the largest WGS study in CyKD to date, we were underpowered to detect common variant signals in our seqGWAS with an OR<3, this has limited our ability to find tractable signals in all forms of GWAS as well as in the heritability analysis which is reliant on this. Furthermore, while WGS allows for more accurate SV calling over traditional microarray, this process is highly dependent on the algorithm used for calling with the possibility of false positives. Ideally long-range sequencing and independent validation would allow for more complete SV detection. Equally, we were unable to functionally characterize the splice variants given the lack of RNA sequencing, meaning our conclusions rest on the enrichment of such variants in cases compared with controls and any clinical actionability for participants in the 100KGP would be subject to cDNA confirmation on a case-by-case basis.

Our study provides the most comprehensive WGS analysis of CyKD to date highlighting the contributions to disease risk of different types of variants across the allelic spectrum. These findings can be used to inform genetic sequencing and counselling strategies offered to patients.

## Methods

### The 100,000 Genomes Project

The 100,000 Genomes Project (100KGP) is one of the largest disease-based sequencing initiatives in the world in which WGS data from large numbers of NHS patients with rare diseases and cancer, and their relatives, have been generated^19^. Key strengths of this dataset with respect to the study of rare diseases are that all germline samples are processed and analysed using a shared pipeline and that sequencing data is available for many individuals without the phenotype under study, drawn from the same population. This allows for robust control of technical artefacts, allele frequency and variant burden in the population, in contrast to previous sequencing studies.

Recruitment to the 100KGP is via a network of 13 NHS Genomic Medicine Centers (GMCs) and includes collection of phenotype data hierarchically encoded using Human Phenotype Ontology (HPO) codes^20^, facilitating computerized analysis of clinical features. CyKD patients were recruited to the project if they met the following criteria:

- >5 cysts affecting one or both kidneys with at least one of the following features:
  - cysts not clinically characteristic of ADPKD;
  - onset before the age of 10;
  - syndromic features;
  - where a genetic diagnosis would influence management;
- and/or:
  - features suggestive of classical ADPKD who had not undergone prior genetic testing of *PKD1* and *PKD2*.

Participants were excluded if they suffered from kidney failure (KF) due to identified (non-cystic) disease, if they had multicystic dysplastic kidney(s) or if they had a prior genetic diagnosis for their condition. We used the Genomics England dataset (v15)^21^, which contains WGS data, details of clinical phenotypes encoded using Human Phenotype Ontology (HPO) terms and structed data automatically extracted from National Health Service (NHS) hospital records, collected for more than 90,259 cancer and rare disease patients (see Data availability) as well as their unaffected relatives to generate the cohorts. Ethical approval for the 100KGP was granted by the Research Ethics Committee for East of England – Cambridge South (REC Ref: 14/EE/1112). Figure 1 details the study workflow, and a full description of the cohort creation can be found in Supplementary Methods 1-4. After quality control, relatedness filtering and ancestry matching (Supplementary Methods 1-4 and Supplementary Figure 1), we were left with 1,209 cases and 26,096 controls for analysis.

Diagnostic variants were ascertained through a combination of the clinical arm of the 100KGP and bioinformatically as detailed in Supplementary Methods 5 to create subgroups stratified by primary molecular diagnosis causing CyKD. We performed all single-variant, gene-burden and structural variant analysis in the total cohort and each molecular subgroup (except the “other genes” group). We used the same controls for each subgroup without repeating ancestry matching as there was no evidence of genomic inflation within each subgroup and the controls (lambda between 0.99-1.02 in all common variant analyses, see Supplementary Figure 7).

### Single-variant seqGWAS

Whole-genome single-variant association analysis (seqGWAS) was carried out using the R package SAIGE^22^ (version 0.42.1) which uses a generalised linear mixed model (GLMM) to account for population stratification. High-quality, autosomal, bi-allelic, LD-pruned SNVs with MAF >5% were used to generate a genetic relationship matrix and fit the null GLMM. Sex and the top 10 principal components were used as covariates (fixed effects). SNVs and indels with MAF ≥0.1% that passed the following quality control filters were retained: minor allele count (MAC) ≥20, missingness <1%, Hardy-Weinberg equilibrium (HWE) p>10^−6^, and differential missingness p>10^−5^. When case-control ratios are unbalanced, as in our study, type 1 error rates are inflated because the asymptotic assumptions of logistic regression are invalidated. SAIGE employs a saddle point approximation^23^ to calibrate score test statistics and obtain more accurate p-values than the normal distribution.

One limitation of SAIGE is that the betas estimated from score tests can be biased at low MACs and therefore odds ratios for variants with MAF <1% were calculated separately using allele counts in R. The R packages qqman^24^ was used to create Manhattan and Q-Q (quantile-quantile) plots. A P-value threshold of 5×10^−8^ for genome wide statistical significance was used for variant-based tests and Bonferroni correction for the number of genes/regions were used for gene or region-based tests. The genomic inflation factor (lambda), calculated based on the 50th percentile, was between 0.99-1.02 in all analyses indicating no significant population stratification.

### Metanalysis of GWAS data

A metanalysis of cystic kidney disease GWAS using summary statistics from our analysis, a combined UK/Japanese Biobank (UKBB/JBB) analysis of 220 phenotypes including polycystic kidney disease (19,093,042 variants) and Finngen (version 8) analysis of cystic kidney disease^25^ (19,441,692 variants) was performed using METAL [version 2011-03-025]^26^. The summary statistics from the UKBB/JBB analyses were lifted over from build 37 to 38 using the UCSC liftover tool^27^. A full breakdown of the biobank phenotypes can be found in Supplementary Methods 6. Between the three data sets 8,217,458 variants were shared with matching alleles. Meta-analysis was performed weighting the effect size estimates using the inverse of the standard errors. Variants showing heterogeneity of effect between the two datasets (PD<D1D×D10^−5^) and those in which the minimum/maximum allele frequencies differed by >0.05 were excluded leaving 6,641,352 variants across 2,923 cases and 900,824 controls. The genomic inflation factor (lambda), calculated based on the 50th percentile, was 1.01 indicating no significant population stratification.

### Single-variant seqGWAS time to event analysis

Genetic Analysis of Time-to-Event phenotypes (GATE)^28^ was used to conduct a time to event (TTE) analysis utilizing the Cox proportional hazard model that accounts for heavily censored phenotypes and low frequency variants. The 100KGP project participants consented to give access to their Hospital Episode Statistics (HES) which is a database containing details of all admission, emergency attendances and outpatient appointments at NHS hospitals in England. The database was searched for codes that would highlight whether a patient had reached kidney failure (KF) as well as codes pertaining to their stage of chronic kidney disease (CKD) from stage 1-5 (Supplementary Table 1). The age of KF was used as the end point in the TTE analysis and those who were yet to reach KF were censored. The same genomic and phenotype data as per the single variant seqGWAS was used to conduct the TTE GWAS.

### Heritability estimation using common variants

Narrow sense heritability (h^2^), the contribution of phenotypic variation from additive genetic factors was estimated using two methods: GCTA-LDMS^29^ and LDAK-SumHer^30^. GCTA-LDMS was applied to the 100KGP WGS data using a European subset of the total ancestry-matched CyKD cohort (full details in Supplementary Methods 7).

Summary statistics from the cystic kidney disease analysis of the combined European ancestry Finngen and UK BioBank (780 FinnGen cases and 424 UKBB cases with 375,708 FinnGen controls and 417,905 UKBB controls) were used with LDAK-SumHer to calculate heritability under the BLD-LDAK model using the pre-computed taggings (a record of the relative expected heritability tagged by various predictors), calculated from the UKBB.

The observed heritability was then liability adjusted to account for the population prevalence of CyKD relative to its representation in the 100KGP^31^. In this analysis a CyKD prevalence of 0.001 was used to transform the observed heritability to a liability threshold model.

### Rare variant collapsing analysis

Prior to collapsing analysis, variants were filtered based on different criteria called masks. The masks used for this analysis were a rare, damaging missense mark (“missense+”), a high confidence loss of function mask (“LoF”), an intronic mask (“intronic”), a splice site mask, a 3-prime untranslated region mask (3’-UTR) and a 5-prime UTR mask (5’-UTR). Full details of the mask parameters and quality control can be found in Supplementary Methods 8.

We applied the “missense” and “LoF” masks to the total cohort and then removed cases that had qualifying variants in statistically significant genes until we had a cohort of patients with “no variants detected” (NVD). To this cohort we applied all the masks listed above looking for previously undetected gene signals. Association testing was performed using the Scalable and Accurate Implementation of Generalized mixed model (SAIGE-GENE) (v0.42.1)^32^ to ascertain whether rare coding variation was enriched in cases on a per-gene basis exome-wide. Sex and the top ten principal components were included as fixed effects when fitting the null model. Full details about the use of SAIGE can be found in Supplementary Methods 9.

A Bonferroni adjusted P-value of 2.58×10^−6^ (0.05/19,364 genes) was used to determine the exome-wide significance threshold. Binary odds ratios and 95% confidence intervals were calculated for exome-wide significance genes by extracting the number of cases and controls carrying qualifying variants per gene in the collapsing analysis and applying a Fisher’s test in R.

### Pathway analysis

For the cohort of patients that had no molecular diagnosis, the summary statistics from their rare variant SKAT-O analysis with SAIGE was analysed using the Gene set analysis Association using Sparse Signals method (GAUSS)^33^ with default settings. The summary statistics were analysed using the canonical (3759 pathways) and hallmark (50 pathways) curated gene set pathways from the Gene Set Enrichment Analysis (GSEA) group^34^. The results of these analyses can be found in Supplementary Table 23-24.

### Exome-wide Structural variant analysis

Structural variants were called from WGS using the Genomics England pipeline that incorporates CANVAS^35^ to detect copy number (>10kb) and MANTA^36^ to identify SVs greater than 50bp. CANVAS uses read depth to assign CNV losses and gain. MANTA uses both discordant read-pair and split-read data to identify SV regions. While MANTA can detect deletions and tandem duplications <10kb, inversions, and interchromosomal translocations, it cannot reliably identify dispersed duplications, small inversions (<200bp), fully assembled large insertions (>2×150bp) or breakends where repeat lengths approach the read size (150bp). Very few insertions were identified in this cohort using MANTA and in view of this they were excluded from downstream analysis. In addition, variants classified as translocations, single breakends or complex SVs which are more difficult to accurately resolve were filtered out.

The following quality control filters were applied to the variants: CNV length > 10kb and Q-score ≥ Q10 indicating 90% confidence there is a variant present, a quality score ≥ 20 indicating 99% confidence that there is a variant at the site, GQ ≥ 15 indicating 95% confidence that the genotype assigned to a sample is correct, and MaxMQ0Frac < 0.4 which indicates the proportion of uniquely mapped reads around either breakend. Variants without paired read support, inconsistent ploidy, or depth > 3× the mean chromosome depth near one or both breakends were excluded.

For each sample BEDTools^37^ was used to extract SVs that intersected at least one exon by a minimum of 1 bp. Variants were then separated by type into CNV, deletion (DEL), duplication (DUP), and inversion (INV) sets before being filtered using BEDTools to remove common SVs of the same type. SVs were removed if they had a minimum 70% reciprocal overlap with the gnomAD SVs^38^ with allele frequency >1% and or a dataset of common (AF>0.1%) SVs generated from 12,243 cancer patients recruited to 100KGP. SVs were then merged using SURVIVOR^39^ allowing a maximum distance of 300bp between pairwise breakpoints and allele frequencies calculated using BCFtools^40^.

After removal of overlapping common variants, a custom Perl script (Dr Helen Griffin, Newcastle University) was used to calculate allele frequencies for each type of SV across the combined case-control cohort using bins of 10kb across the entire genome. SVs with an AF < 0.1% were retained for further analysis.

Exome-wide gene-based burden testing was carried out using custom R scripts stratified by SV type. SVs were aggregated across 19,005 autosomal protein-coding genes. A two-sided Fisher’s exact test was used to compare SVs in cases and controls under a dominant inheritance model. The Bonferroni correction for the number of genes (*P=*0.05/19,005=2.6×10^−6^) tested was applied, although with the knowledge that this is likely to be too stringent given the tests are not truly independent (one SV can affect multiple genes).

### Power

PAGEANT^41^ is a power calculation tool for rare variant collapsing tests that uses the underlying distribution of gene size and MAF of variants from the ExAC dataset^42^. Under the assumption that 80% of variants collapsed per gene in the SAIGE-GENE analysis were causal, we calculated that we had >80% power to detect a gene signal that accounted for >4% of the variance of the phenotype with an exome-wide significance threshold of *P=*2.5×10^−6^ in the total case control cohort.

For single variant analysis, power was calculated using the R package genpwr^43^ under an additive model using the conventional genome-wide significance threshold of *P<*5×10^−8^. With this sample size at an allele frequency of 1%, single variant association testing was sufficiently powered (> 80%) to detect alleles with an odds ratio (OR) > 3.

For heritability analysis we used the GCTA-GREML power calculator^44^ which revealed we had a 54% chance of detecting 5% heritability within the 100KGP cohort. This is likely to be even lower in the summary statistics methods applied to the combined UKBB/FinnGen metanalysis.

## Data Availability

All collapsing gene analyses can be found in the Supplementary Tables.

All seqGWAS summary statistics can be found at: https://zenodo.org/records/10613736

Details of the aggregated dataset used for the analysis can be found at: https://re-docs.genomicsengland.co.uk/aggv2/

Genomic and phenotype data from participants recruited to the 100,000 Genomes Project can be accessed by application to Genomics England Ltd (https://www.genomicsengland.co.uk/about-gecip/joining-research-community/).

## Code Availability

Code used for the analyses in this paper can be found at: https://github.com/oalavijeh/cykd_paper.

Details of the rare variant workflow can be found at: https://re-docs.genomicsengland.co.uk/avt/

Details of the common variant GWAS workflow can be found at: https://re-docs.genomicsengland.co.uk/gwas/

## Supporting information

supplementary methods and images

## Data Availability

All data produced in the present work are contained in the manuscript and online at https://zenodo.org/records/10613736

https://github.com/oalavijeh/cykd_paper

## Acknowledgments

This research was made possible through access to the data and findings generated by the 100,000 Genomes Project. The 100,000 Genomes Project is managed by Genomics England Limited (a wholly owned company of the Department of Health and Social Care). The 100,000 Genomes Project is funded by the National Institute for Health Research and NHS England. The Wellcome Trust, Cancer Research UK and the Medical Research Council have also funded research infrastructure. The 100,000 Genomes Project uses data provided by patients and collected by the National Health Service as part of their care and support. The authors gratefully acknowledge the participation of the patients and their families recruited to the 100,000 Genomes Project.

OSA is funded by an MRC Clinical Research Training Fellowship (MR/S021329/1). MYC is funded by a Kidney Research UK Clinical Research Fellowship (TF_004_20161125). DPG is supported by the St Peter’s Trust for Kidney, Bladder and Prostate Research.

## Author Contributions

DPG conceived the study. OSA conducted the analyses and wrote the manuscript with extensive input from MYC, DPG and APL. GD aided with analyses; CV provided scripting support. AS, AK and AH provided bioinformatic assistance from the Genomics England side. HS, DB, RS, APL and DPG provided tutelage and guidance throughout.

## Figures

Figure 1 – Study workflow highlighting the creation of the cohort and analysis steps.

Figure 2 – Kaplan-Meier plot of kidney survival plotted by primary driving variant.

Figure 3 – Rare variant collapsing analysis under the missense+ mask.

Figure 4 – Rare variant collapsing analysis under loss-of-function mask.

Figure 5 – Gene based Manhattan plot of the SAIGE-GENE analysis with the splice mask

Figure 6 – Gene based Manhattan plot of the exome-wide gene based structural variant analysis

Figure 7 – Manhattan plot of CyKD metanalysis.

## Tables

Table 1 – Demographic and phenotypic breakdown of the recruited cystic kidney disease probands and controls

Table 2 – Molecular diagnosis in cystic kidney disease cases that were solved by the 100,000-genomes project clinical pipeline.

Table 3 – Cohort breakdown by variant type in the 100KGP CyKD cohort after clinical and research pipeline

Table 4 – Odds ratio of developing CyKD in the 100KGP (n=741) and the UKBB (n=825) in each different gene.

