## supplementary methods and images for "Quantifying variant contributions in cystic kidney disease using national-scale whole genome sequencing"

### Contribution of variants across the allelic frequency spectrum to cystic kidney disease. - Supplementary appendix and figures

#### Contents

**Members of the Genomics England Research Consortium**

**Supplementary Methods**

Supplementary Methods 1: Cohort creation and rational

Supplementary Methods 2: Genomic variant calling format file annotation and variant-level quality control

Supplementary Methods 3: Relatedness estimation and principal components analysis

Supplementary Methods 4: Ancestry-matching of cases and controls

Supplementary Methods 5: Subgroup analysis stratified by primary variant and depleting analysis

Supplementary Methods 6: Other biobanks used for meta-analysis.

Supplementary Methods 7: Heritability using common variants.

Supplementary Methods 8: Rare variant selection for collapsing analysis.

Supplementary Methods 9: Background to using SAIGE-GENE.

#### Figures

Supplementary Figure S1 - Ancestry matching.
Supplementary Figure S2 Rare variant analysis of the no variant detected cohort.
Supplementary Figure S3 – 100KGP CyKD GWAS
Supplementary Figure S4 – Finngen CyKD GWAS
Supplementary Figure S5 – UKBB/JBB CyKD GWAS
Supplementary Figure S6 - Manhattan plots of GWAS by primary driving variant
Supplementary Figure S7 - QQ plots of primary variant GWAS
Supplementary Figure S8 - Manhattan plots of time-to-event GWAS in the total CyKD and primary driving variant cohorts.
Supplementary Figure S9 – Power calculations for the CyKD GWAS

#### References

### Members of the Genomics England Research Consortium

**Genomics England Research Consortium**

John C. Ambrose[1]; Prabhu Arumugam [1]; Roel Bevers [1]; Marta Bleda [1]; Freya Boardman-Pretty [1],2; Christopher R. Boustred [1]; Helen Brittain [1]; Mark J. Caulfield [1,2]; Georgia C. Chan [1]; Greg Elgar [1, 2]; Tom Fowler [1]; Adam Giess [1]; Angela Hamblin [1]; Shirley Henderson [1],2; Tim J. P. Hubbard [1]; Rob Jackson [1]; Louise J. Jones [1],2; Dalia Kasperaviciute [1, 2]; Melis Kayikci [1]; Athanasios Kousathanas [1]; Lea Lahnstein [1]; Sarah E. A. Leigh [1]; Ivonne U. S. Leong [1]; Javier F. Lopez [1]; Fiona Maleady-Crowe [1]; Meriel McEntagart [1]; Federico Minneci [1]; LoukasMoutsianas [1, 2]; Michael Mueller [1, 2]; Nirupa Murugaesu [1]; Anna C.Need [1, 2]; Peter O'Donovan [1]; Chris A.Odhams [1]; Christine Patch [1, 2]; Mariana Buongermino Pereira [1]; Daniel PerezGil [1]; John Pullinger [1]; Tahrima Rahim [1]; Augusto Rendon [1]; Tim Rogers [1]; Kevin Savage [1]; Kushmita Sawant [1]; Richard H. Scott [1]; Afshan Siddiq [1]; Alexander Sieghart [1]; Samuel C. Smith [1]; Alona Sosinsky [1, 2]; Alexander Stuckey [1]; Mélanie Tanguy [1]; Ana Lisa Taylor Tavares [1]; Ellen R. A. Thomas [1],2; Simon R. Thompson [1]; Arianna Tucci [1, 2]; Matthew J. Welland [1]; Eleanor Williams [1]; Katarzyna Witkowska [1, 2]; Suzanne M.Wood [1, 2].

1. Genomics England, London, UK

2. William Harvey Research Institute, Queen Mary University of London, London, EC1M 6BQ, UK.

### Supplementary Methods

#### Supplementary Methods 1: Cohort creation and rational

Cases were included if they were recruited under the primary diagnosis of "cystic kidney disease". All cases recruited had been assessed in the clinical interpretation arm of the 100KGP. This involved ascertainment of variants in an expert curated panel of 28 cystic genes within PanelApp (https://nhsgms-panelapp.genomicsengland.co.uk/panels/283/v4.0) with multi-disciplinary review and application of American College of Molecular Genetics (ACMG) criteria to determine pathogenicity[1]. The control cohort consisted of 27,660 unaffected relatives of non-renal rare disease participants, excluding those with HPO terms and/or hospital episode statistics (HES) data consistent with secondary causes of kidney disease or kidney failure. By utilizing a case-control cohort sequenced on the same platform, we aimed to minimize confounding by technical artefacts.

Whole genome sequencing was performed by Genomics England, as described previously[2].

##### Cohort rationale

Given the small number of recruited cases, we chose to jointly analyze individuals from diverse ancestral backgrounds, thereby preserving sample size and boosting power. To mitigate confounding due to population structure whilst using this mixed ancestry approach we employed two strategies, as previously described3. First, we carried out ancestry-matching of cases and controls using weighted principal components and second, utilized a generalized logistic mixed model to account for relatedness between individuals; further details can be found in supplementary methods 3.

#### Supplementary Methods 2: Genomic variant calling format file annotation and variant-level quality control

All genomic files were aligned to the GRCh38 reference human genome25. Genomic variant call format files (gVCFs) were aggregated using gvcfgenotyper (Illumina, version: 2019.02.26) with variants normalized and multi-allelic variants decomposed using vt26 (version 0.57721). Variants were retained if they passed the following filters: missingness ≤ 5%, median depth ≥ 10, median GQ ≥ 15, percentage of heterozygous calls not showing significant allele imbalance for reads supporting the reference and alternate alleles (ABratio) ≥ 25%, percentage of complete sites (completeGTRatio) ≥ 50% and P value for deviations from Hardy-Weinberg equilibrium (HWE) in unrelated samples of inferred European ancestry ≥ 1×10-5. Male and female subsets were analyzed separately for sex chromosome quality control. Per-variant minor allele count (MAC) was calculated across the case-control cohort, MAC is defined as the number of minor alleles counted for each marker. Annotation was performed using Variant Effect Predictor27 (VEP, version 98.2) including CADD28 (version 1.5), and allele frequencies from publicly available databases including gnomAD29 (version 3) and TOPMed30 (Freeze 5). Variants were filtered using bcftools31 (version 1.11).

#### Supplementary Methods 3: Relatedness estimation and principal components analysis

A set of 127,747 high quality autosomal LD-pruned biallelic single nucleotide variants (SNVs) with a minor allele frequency (MAF) > 1% was generated using PLINK[3] (v1.9), MAF is defined as the frequency at which the second most common allele occurs in a given population. SNVs were included if they met all the following criteria: missingness < 1%, median GQ ≥ 30, median depth ≥ 30, AB Ratio ≥ 0.9, completeness ≥ 0.9. Ambiguous SNVs (AC or GT) and those in a region of long-range high LD (Linkage Disequilibrium) were excluded. LD pruning was carried out using an $r^2$ threshold of 0.1 and window of 500kb. SNVs out of HWE in any of the AFR, EAS, EUR or SAS 1000 Genomes populations were removed (pHWE < 1 ×10-5). Using this variant set, a pairwise kinship matrix was generated using the PLINK2 implementation of the KING-Robust algorithm[4] and a subset of unrelated samples was ascertained using a kinship coefficient threshold of 0.0884 (2nd degree relationships). Ten principal components were generated using PLINK2 for ancestry-matching and as covariates in the association analyses.

#### Supplementary Methods 4: Ancestry-matching of cases and controls

Given the mixed-ancestry composition of the cohort we employed a case-control ancestry-matching algorithm to optimize genomic similarity and minimize the effects of population structure as previously described[5] with each case having to match a minimum of two controls to be included in the final cohort. 1209 cases and 26096 controls remained for analysis (Supplementary Figure S1).

#### Supplementary Methods 5: Subgroup analysis stratified by primary variant and depleting analysis

Patients who have their phenotype "solved" by the clinical multi-disciplinary team (MDT) had a report issued with the details of the molecular diagnosis. Depending on the diagnosis these patients were placed into different cohorts: PKD1-truncating (PKD1-T), PKD1-non truncating (PKD1-NT), PKD2-truncating (PKD2-T), PKD2 non-truncating (PKD2-NT), "other gene" (encompassing other genes in the panel) and no variant detected (NVD). In the patients with NVD we bioinformatically reanalyzed them looking for variants that met the "missense+" or "loss-of-function mask" (detailed below), in the approved cystic kidney disease panel of genes in PanelApp[6] and placing them in the relevant cohort. The filtering was performed using bcftools and filter-VEP. For each subsequent round of analysis (across SNV and SVs) if a gene or SV was found to be significantly enriched in cases, we identified the cases that contained qualifying variants and removed them from the NVD cohort and re-analysed the cohort, eventually leaving 184 cases with no clear genetic cause of disease identified.

We performed all single-variant, gene-burden and structural variant analysis in each molecular subgroup (bar the "other genes" group). We used the same controls for each subgroup without repeating ancestry matching as there was no evidence of genomic inflation within each subgroup and the controls (lambda between 0.99-1.02 in all common variant analyses, see supplementary data).

#### Supplementary Methods 6: Other biobanks used for meta-analysis.

All other biobanking studies used imputed datasets and various genotyping platforms which are detailed in their respective publications[7,8]. The Japanese Biobank (JBB) took 510 cases and 178,216 controls recruited from 12 medical institutions in Japan of predominant Japanese ancestry with a diagnostic tag of "polycystic kidney disease". GWAS was conducted in this cohort using SAIGE (v.0.37) using the top 20 principal components and age. The UK Biobank (UKBB) analysis consisted of 424 cases and 355,431 controls matched to the JBB cohort via phecodes, the association study was conducted using SAIGE and the same principal components. The FinnGen analysis consisted of 780 cases and 341,081 controls, cases were recruited if they had the ICD-10 code for cystic kidney disease (Q61) with the association analysis being conducted with REGENIE[9].

#### Supplementary Methods 7: Heritability using common variants.

GCTA-LDMS was applied to the 100KGP WGS data using a European subset of the total ancestry-matched CyKD cohort. Variants with MAF ≥ 0.1% were included. Using the GREML-LDMS approach variants were stratified into seven different bins based on MAF (0.001-0.01, 0.01-0.05, 0.05-0.1, 0.1-0.2, 0.2-0.3, 0.3- 0.4, 0.4-0.5) and for each bin of variants, SNP-based LD scores were calculated over a 200kb region (with 100kb overlap between two adjacent segments). For a given bin of variants defined by MAF, variants were further stratified into quartiles using LD scores. For each of the 28 bins subset by MAF and LD, GCTA was used to produce a GRM from the raw genotype files. The REML (restricted maximum likelihood) function was then used to conduct a GREML-LDMS analysis using the 28 GRMs, including the top four principal components as covariates. GREML has not been validated for mixed ancestry cohorts, so an estimation was made using a European subset of the CyKD cohort as defined by principal component analysis (903 cases and 20255 controls).

Summary statistics from the cystic kidney disease analysis of the combined European ancestry Finngen and UK biobank (865 FinnGen cases, 505 UKBB cases, 375708 FinnGen controls and 417905 UKBB controls) were used with LDAK-SumHer to calculate heritability under the BLD-LDAK model using the pre-computed taggings, calculated from the UK Biobank.

The observed heritability was then liability adjusted to account for the population prevalence of CyKD relative to its representation in the 100KGP[10]. In this analysis a CyKD prevalence of 0.001 was used to transform the observed heritability to a liability threshold model.

#### Supplementary Methods 8: Rare variant selection for collapsing analysis.

Single variant association testing is underpowered when variants are rare and a collapsing approach which aggregates variants by gene or genomic region can be adopted to boost power. Whilst the variants are aggregated by gene, they are also filtered to help boost power. Each set of filters are applied as a "mask" which instructs the workflow to collapse variants as per a set of parameters. The mask used for this analysis were a rare, damaging missense mark ("missense+"), a high confidence loss of function mask ("LoF"), an intronic mask ("intronic"), a splice site mask, a 3-prime untranslated region mask (3'-UTR) and a 5-primte UTR mask (5'-UTR).

For the missense+ mask we extracted coding SNVs and indels with MAF < 0.01% in gnomAD annotated with one of the following: missense, in-frame insertion, in-frame deletion, start loss, stop gain, frameshift, splice donor, splice acceptor for each gene and further filtered them by CADD[11] (v1.5) score using a threshold of ≥20 corresponding to the top 1% of all predicted deleterious variants in the genome. Variants meeting the following quality control filters were retained: MAC ≤ 20, median site-wide sequencing depth in non-missing samples > 20 and median GQ ≥ 30. Sample-level QC metrics for each site were set to minimum depth per sample of 10, minimum GQ per sample of 20 and ABratio P value > 0.001.

Variants with significantly different missingness between cases and controls (P<10-5) or >5% missingness overall were excluded. For the dedicated loss-of-function (LoF) analysis, variants were selected that were deemed to be of "high confidence" (HC) by the loss-of-function transcript effect estimator (LOFTEE)[12]. These assessed variants are stop-gained, splice site disrupting or frameshifts and collapsed on a per-gene basis. The same quality controls were applied as above. For the splice site mask, variants were collapsed if their SpliceAI scores were greater than 0.8 (on a scale of 0-1) indicating variants that were highly likely to affect splicing. These were divided by their predicted effect on splicing (donor loss, donor gain, acceptor loss and acceptor gain). The intronic mask was defined by variants labelled as "intronic" by VEP with a CADD score greater than 20 and with a MAF<0.01% in gnomAD. The 5' and 3' UTR masks were defined by variants with a MAF<0.01% in gnomAD, a CADD score greater than 10 and found within their respective UTR region as labelled by VEP.

#### Supplementary Methods 9: Background to using SAIGE-GENE.

SAIGE-GENE uses a generalized mixed-model to correct for population stratification and cryptic relatedness as well as a saddle point approximation and efficient resampling adjustment to account for the inflated type 1 error rates seen with unbalanced case-control ratios. It combines single-variant score statistics and their covariance estimate to perform SKAT-O34 gene-based association testing, upweighting rarer variants using the beta (1,25) weights option. SKAT-O is a combination of a traditional burden and variance-component test and provides robust power when the underlying genetic architecture is unknown.

1a

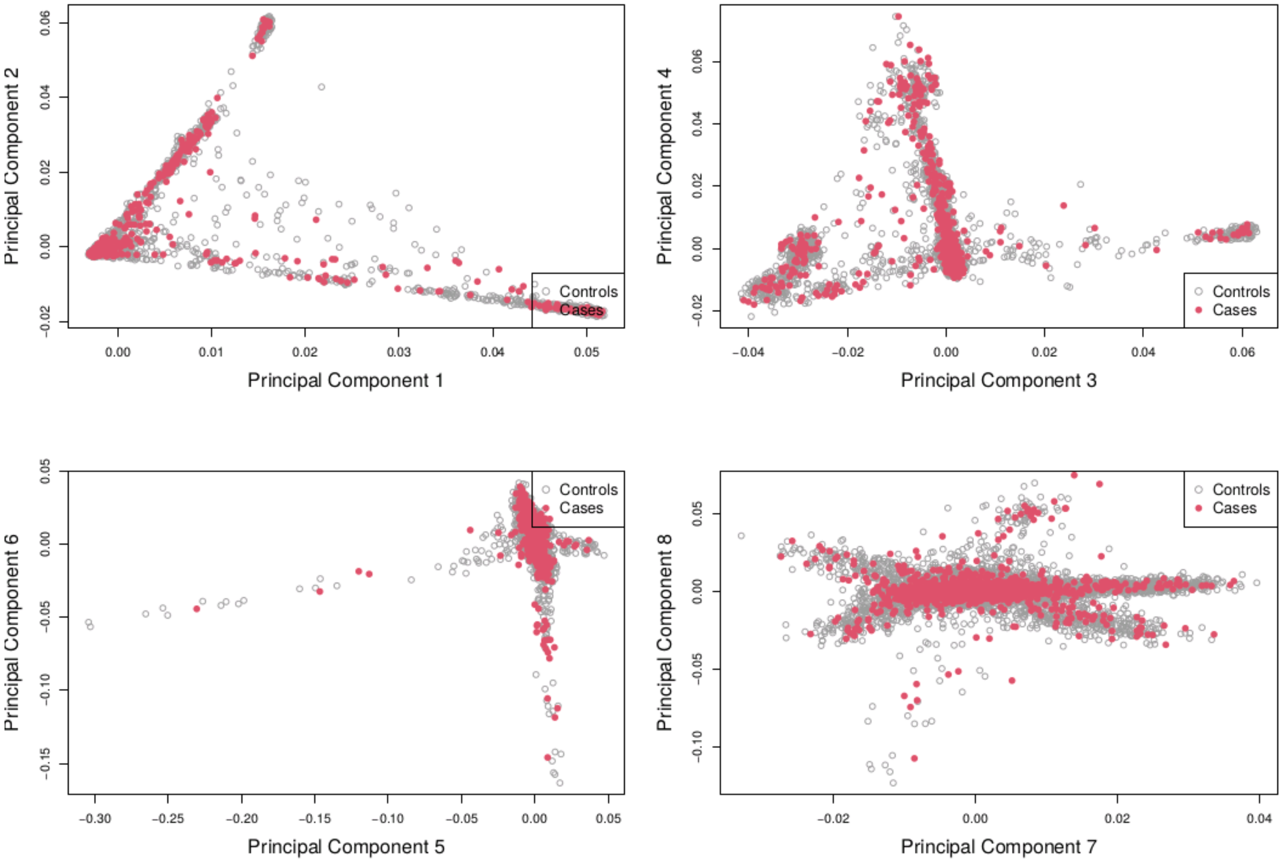


1b

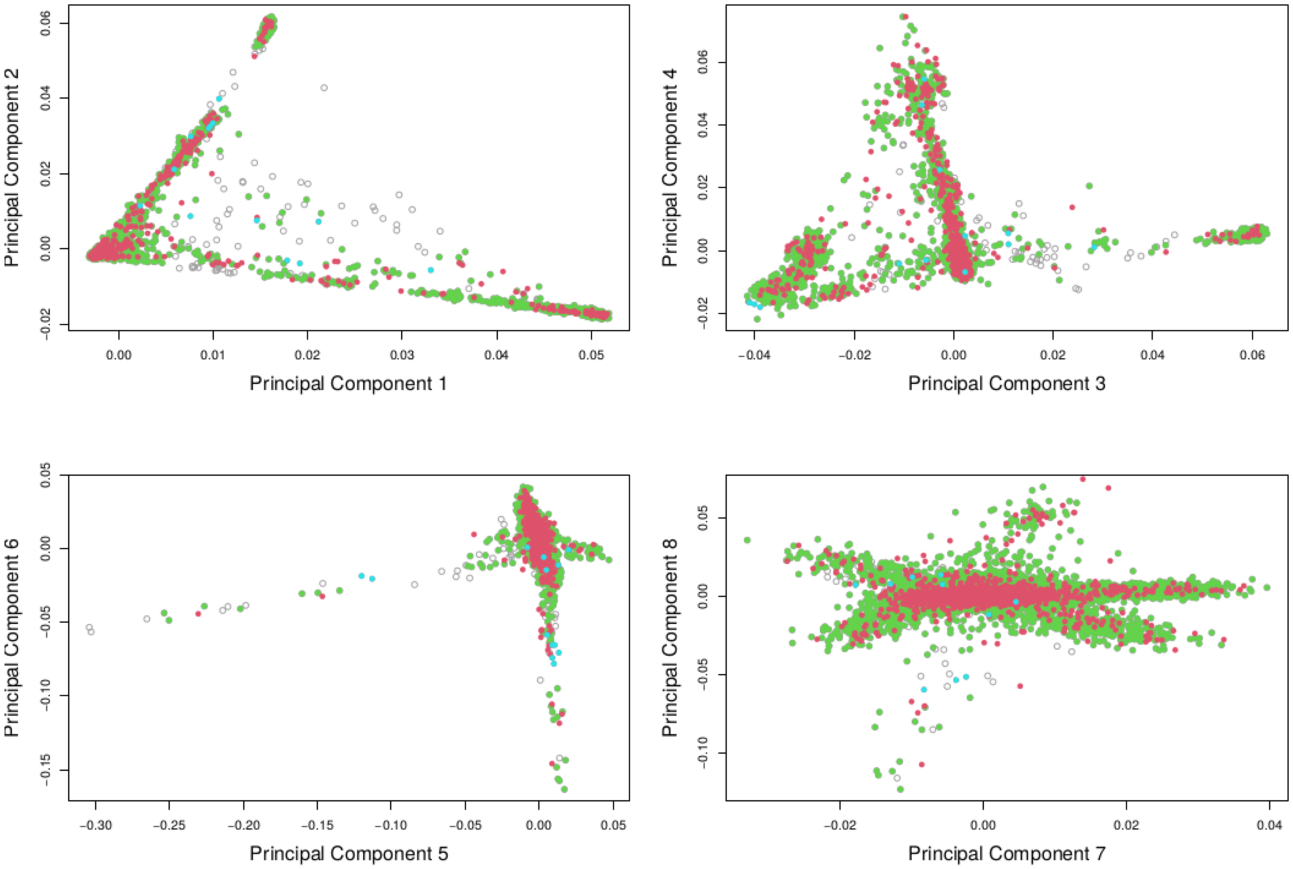


*Supplementary Figure S1 - Ancestry matching.*

*1a. Principal component analysis showing the first eight principal components for matched cases (red) and controls (white). 1b. Principal component analysis showing the first eight principal components for matched cases (red) and controls (green) and unmatched controls (grey). This highlights that cases are taken from multiple different ancestries with the appropriate matched controls.*

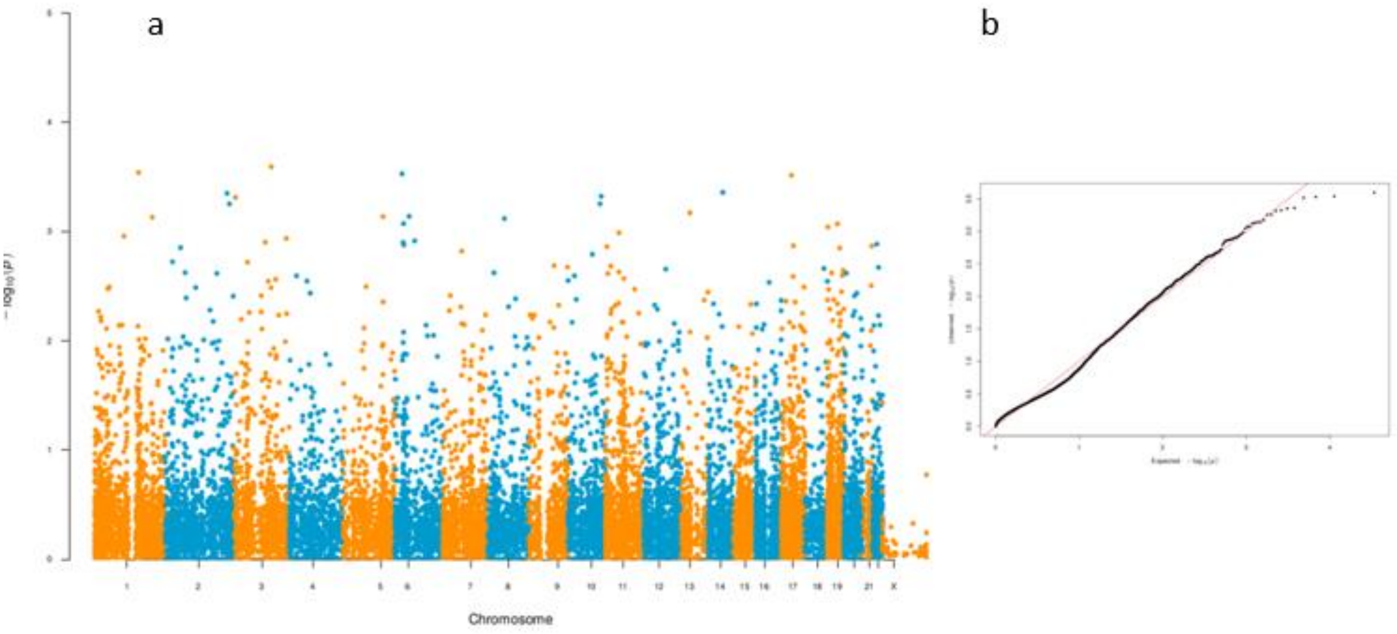


*Supplementary Figure S2 Rare variant analysis of the no variant detected cohort.*

*2a Rare variant analysis of the no variant detected cohort depleted for COL4A3 and IFT140 cases including 266 cases and 26096 controls under the “missense+” mask showing no enrichment of genes. 2b - quantile-quantile plot of the rare variant burden analysis of 266 cases and 26096 controls under the “missense+” mask.*

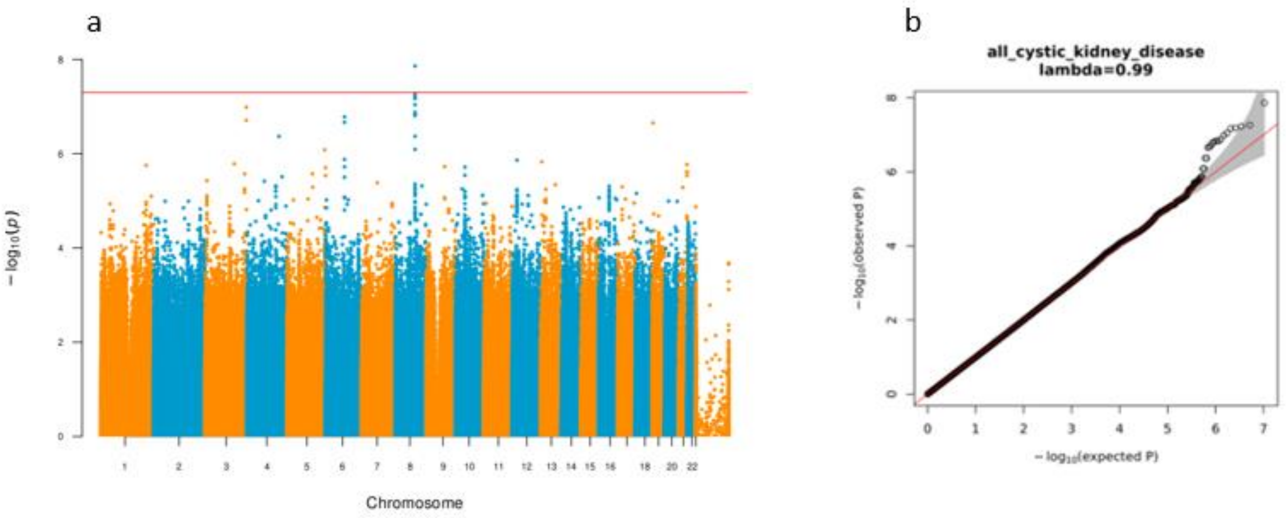


*Supplementary Figure S3 – 100KGP CyKD GWAS*

*3a seqGWAS Manhattan plot of 1209 cystic kidney disease cases against 26096 ancestry matched controls. There is no variant that reaches genome wide significance. 3b – Quantile-Quantile plot of the association test in 3a. The genomic inflation is 0.99.*

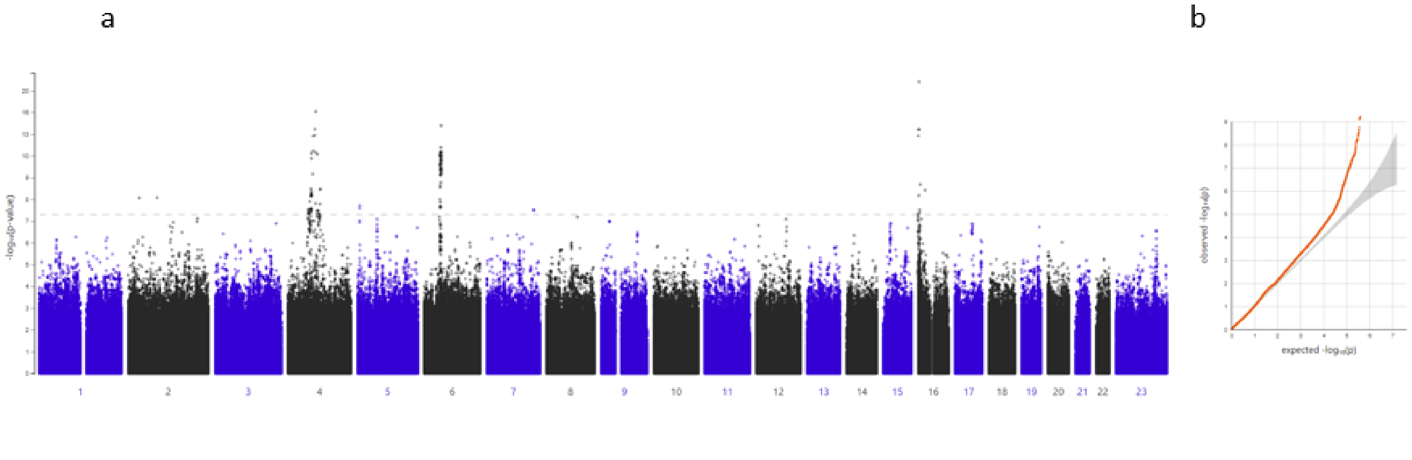


*Supplementary Figure S4 – Finngen CyKD GWAS*

*4a GWAS Manhattan from Finngen plot of 780 cystic kidney disease cases against 375708 controls. 4b – Quantile-Quantile plot of the association test in 3a. The genomic inflation is 1.04.*

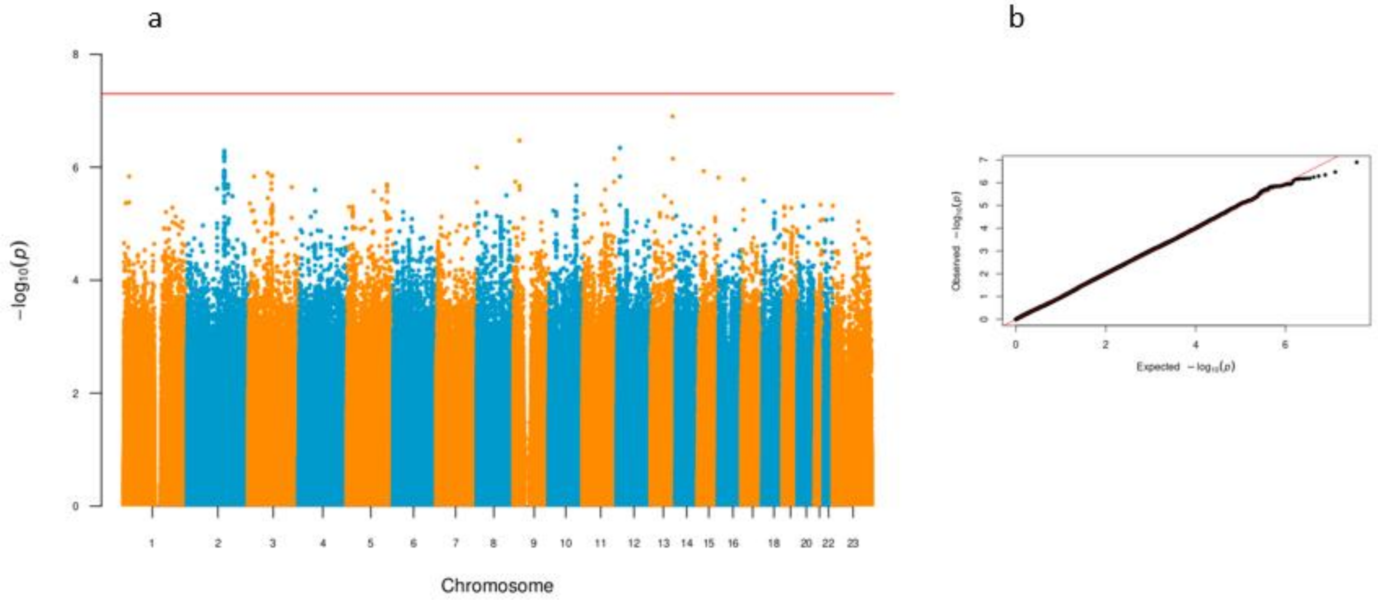


*Supplementary Figure S5 – UKBB/JBB CyKD GWAS*

*5a GWAS Manhattan from UKBB/JBB analysis - plot of 932 cystic kidney disease cases against 534581 controls. 5b – Quantile-Quantile plot of the association test in 5a. The genomic inflation is 1.02.*

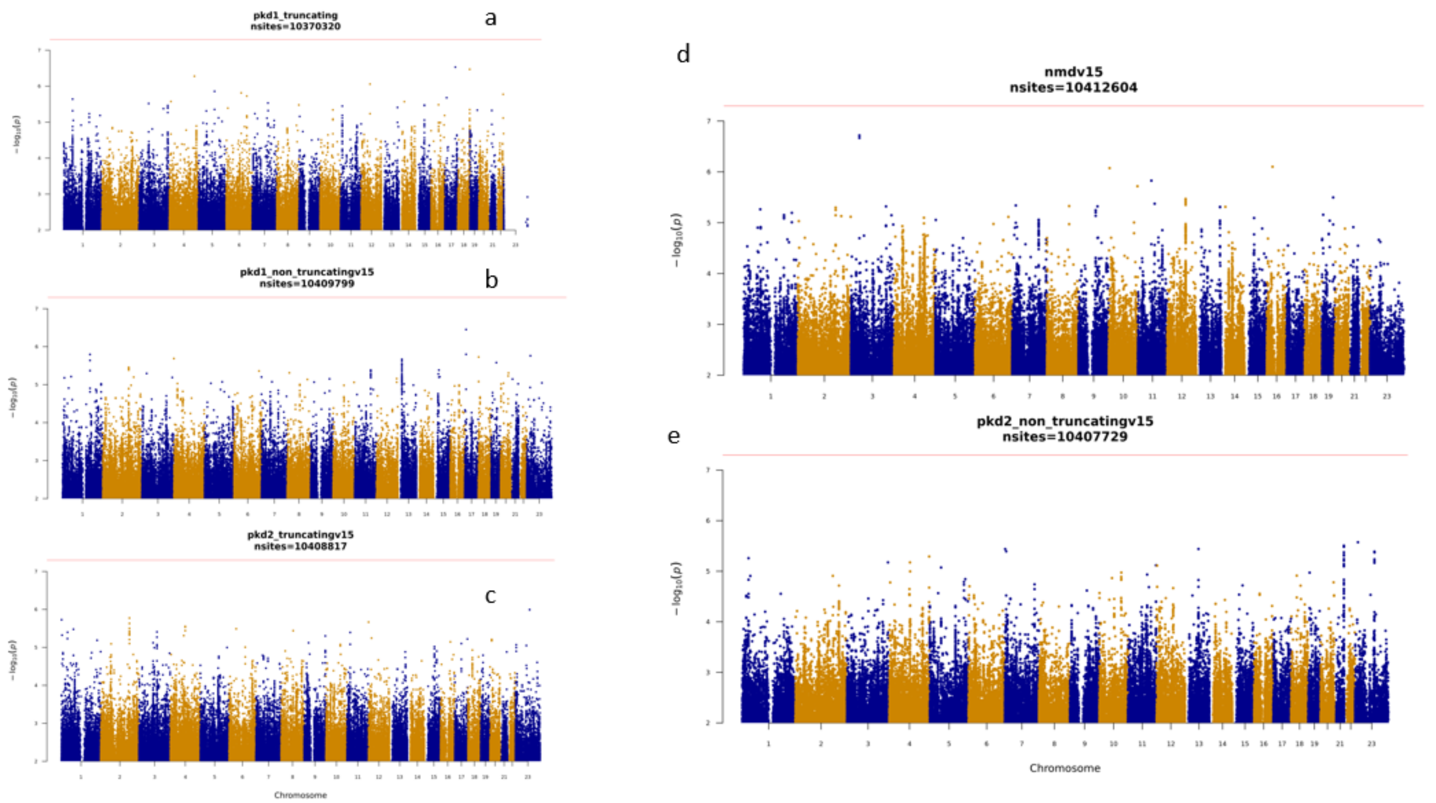


*Supplementary Figure S6 - Manhattan plots of GWAS by primary driving variant*

*SeqGWAS Manhattan plot of cystic kidney disease cases against 26096 ancestry matched controls by primary variant type. 6a – PKD1 truncating, 6b – PKD1 non truncating, 6c - PKD2 truncating, 6d - No primary variant detected, 6e – PKD2 non truncating. There is no variant that reaches genome wide significance in any analysis.*

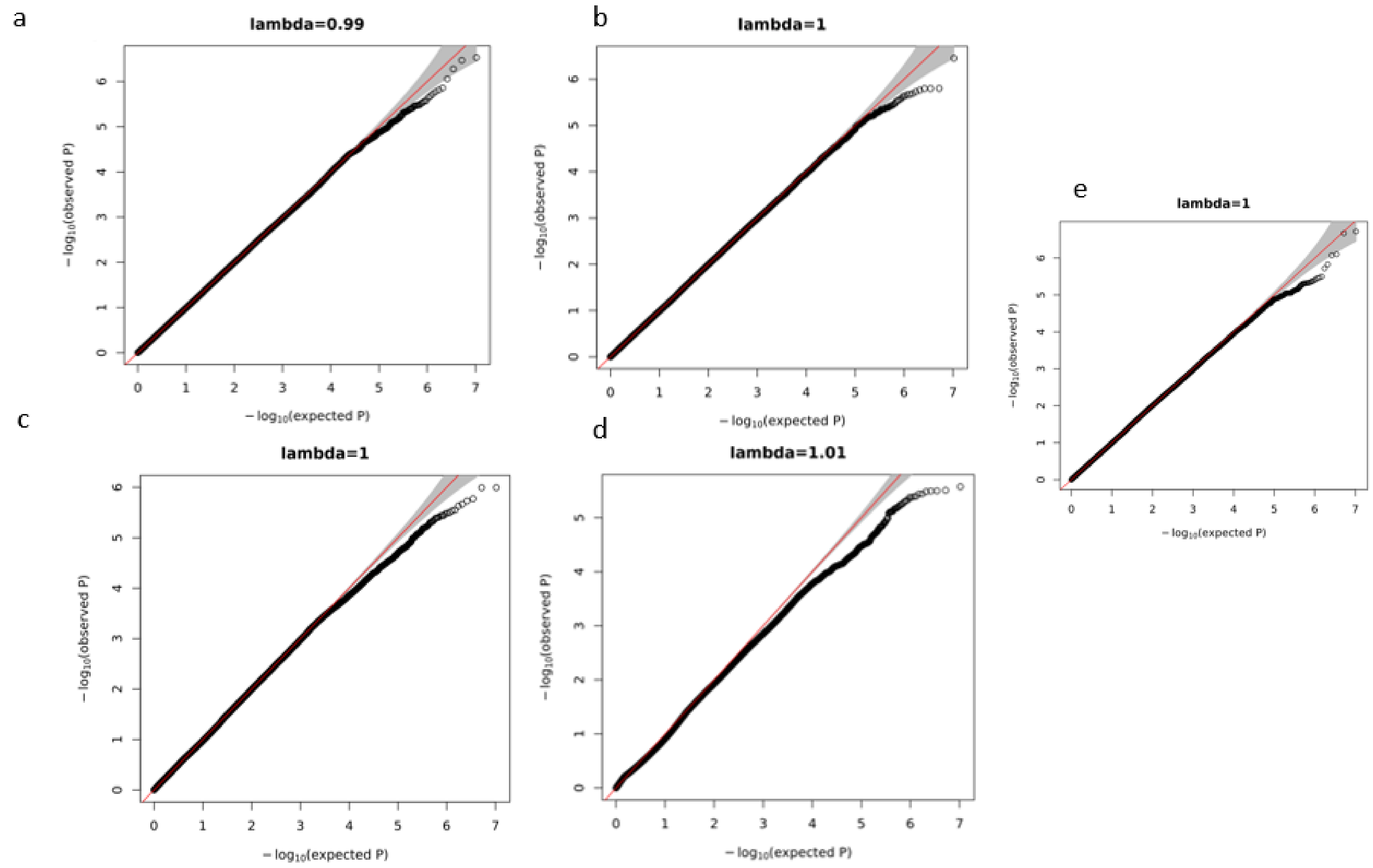


*Supplementary Figure S7 - QQ plots of primary variant GWAS*

*QQ plots SeqGWAS per primary 7a – PKD1 truncating, 7b – PKD1 non truncating, 7c - PKD2 truncating, 7d -PKD2 non truncating, 7e – No primary variant detected. There is no evidence of genomic inflation in the mixed ancestry cohort.*

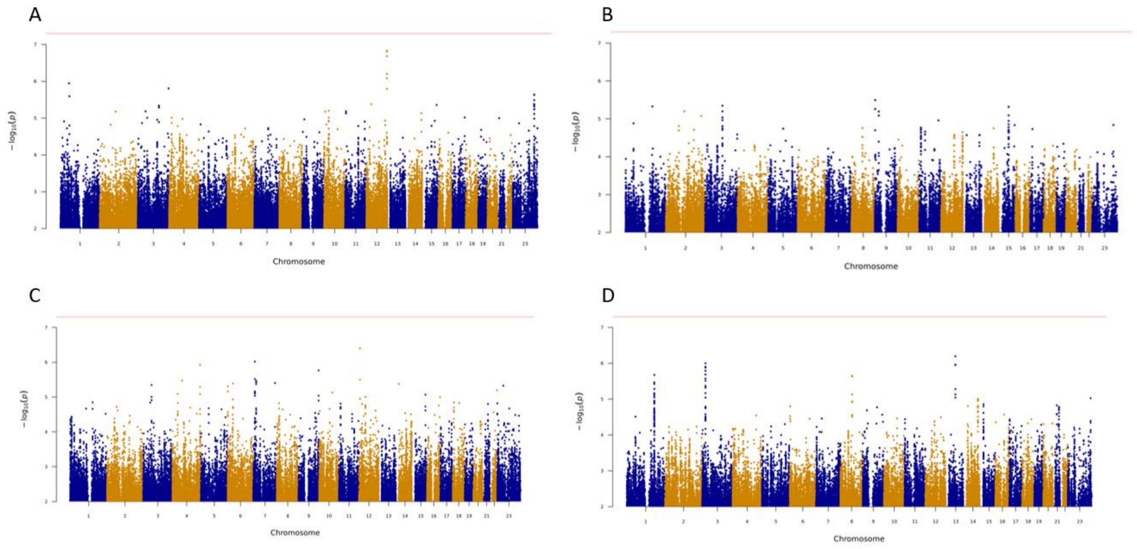


*Supplementary Figure S8 - Manhattan plots of time-to-event GWAS in the total CyKD and primary driving variant cohorts.*

*Manhattan plot of the time-to-event GWAS across the total CyKD cohort and divided by primary variant and no variant detected cohorts. A – Total CyKD cohort (11485299 markers), B – No variant detected cohort (7718522 markers), C – PKD1-truncating cohort (8543817 markers), D – PKD1-nontruncating cohort (7465234 markers). The red line represents genome wide significance. There were too few events for PKD2 to be analysed.*

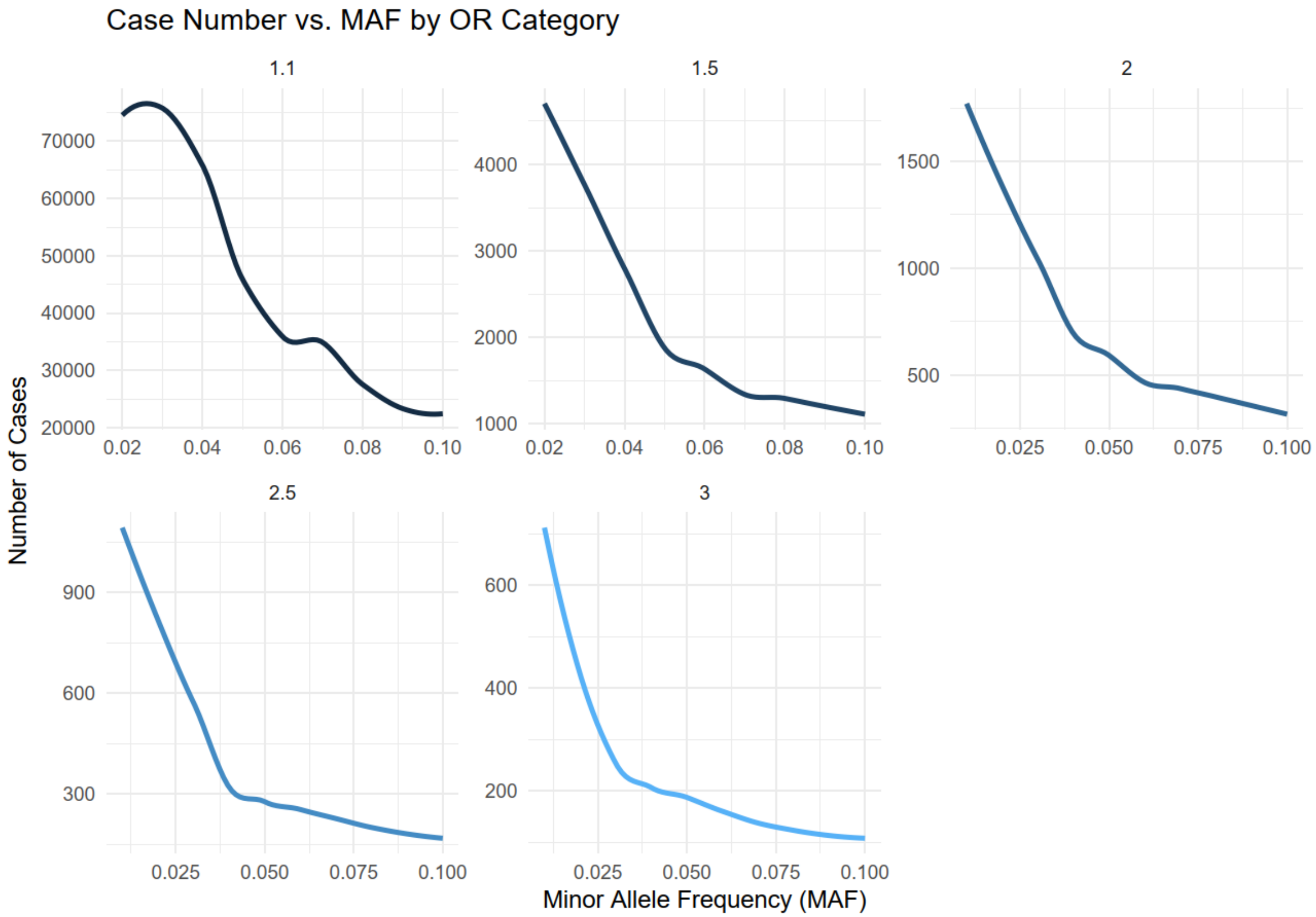


*Supplementary Figure S9 – Power calculations for the CyKD GWAS*

*Plot representing the case numbers against the MAF per OR category to detect a signal in a CyKD GWAS assuming a case rate of 1:1000 at a power of 80% with a genome wide significance of $5x10^{-08}$ under an additive model.*
